# Hepatitis B Prophylaxis Rarely Prevents Subclinical Molecularly Evident HBV Infection in Children Born to HBsAg-Positive Mothers

**DOI:** 10.1101/2025.08.18.25333933

**Authors:** Liang Xu, Qinghai Dai, Huiying Yang, Rui Su, Yanli Shi, Jing Chen, Jing Hao, Jinyu Hao, Wei Lu, Tomasz I. Michalak

## Abstract

Hepatitis B virus (HBV) remains one of the most life-threatening human pathogens causing chronic hepatitis B and hepatocellular carcinoma in millions globally. Protective vaccines are available for decades, but their implementation for infant vaccination has not yet been universally adopted. Mother-to-child transmission remains the prevailing route of HBV spread. Regardless of antiviral treatment of HBV-infected pregnant woman and neonatal HBV immunoprophylaxis, HBV is transferred at low levels to newborns as a subclinical (occult) infection at rates ranging from singular cases to the majority of the infant population examined in different centers worldwide. In the current study of 13 mother-child pairs, we applied a high-sensitivity testing approach validated in our previous studies, examination of different compartments of HBV occurrence and testing of serial samples from infants followed for up to two years after birth to HBV-infected mothers treated or not with antiviral agent before parturition. All newborns received standard HBV immunoprophylaxis with HBV vaccine and hepatitis B immunoglobulin supplemented with additional doses of both during follow-up. The data showed that the majority (84.6%) of the children investigated remained HBV reactive during the 24-month follow-up after birth at levels of <100 HBV genome copies per microgram of DNA, indicating that the prophylactic protocol was only marginally effective in preventing occult infection in newborns. Nonetheless, the same protocol inhibited development of overt HBV infection for at least two years after birth. In addition, the sequencing of the 360-bp HBV preS-S genome fragment revealed homology between infants and umbilical cord blood mononuclear cells that diverged from the corresponding sequences in plasma or maternal peripheral blood cells in 4 of the 5 mother-child pairs examined. These data from the single-center study indicate that occult HBV infection in infants born to HBV-infected mothers is much more frequent than that reported and argue that virus transmission to newborns could be an inevitable consequence of maternal infection with HBV. This study also uncovered previously unknown HBV genomic discrepancy between infected mothers and their newborns with occult infection suggesting the fetal and/or placental origin of the viral variant detected.

## Introduction

An estimated 254 million people have chronic hepatitis B virus (HBV) infection seropositive for HBsAg (HBV surface antigen) [1]. Although safe and effective vaccines are available, HBV remains a serious global health burden with the annual rates of about 1.2 million of new symptomatic infections and up to 1.1 million of deaths mostly due to liver failure, cirrhosis and hepatocellular carcinoma (HCC) [1]. In addition to the HBsAg-positive overt infection, persistent occult HBV infection (OBI) occurs with the expected global incidence of up ten-fold greater than that of the overt infection. OBI is diagnosed when HBV DNA is present in the absence of serum HBsAg detectable by the conventional clinical laboratory tests [2,3].

This clinically silent infection became to a greater extent apparent when polymerase chain reaction (PCR)-based assays were introduced for HBV DNA detection [2,4,5]. OBI is subdivided into seropositive and seronegative based on the presence or not of antibodies to HBV antigens, particularly those directed against HBV core (nucleocapsid) (anti-HBc) [2,3,6].

The virological and pathogenic implications of OBI include risks of HBV transmission *via* blood transfusions and organ, tissue or cell donations, reactivation of hepatitis B due to immunomodulatory and cytotoxic therapies, acceleration of the progression of coinciding liver diseases, and HCC development [7–11].

The most frequent route of HBV spread remains mother-to-child transmission (MTCT). Blocking of this passageway is considered to be the key in limiting the virus perinatal transmission and the likelihood of chronic hepatitis B (CHB) in children born to chronically HBV-infected women [12]. In this regard, both treatment of HBsAg-positive mothers with an anti-HBV directly acting agent (DAA), and HBV vaccination (HepBvac) and passive immunization with hepatitis B immunoglobulin (HBIG) of infants within 12 hours of birth, followed by HepBvac boosters at one and 6 months of age, are recommended [12,13].

However, OBI was reported in the newborns worldwide regardless of the maternal and perinatal HBV prophylaxis in ranges from 3.1% to 64% when tested by assays with varied HBV DNA detection sensitivity [14–20]. The mechanism of vertically acquired OBI in the context of maternal anti-HBV therapy and neonatal HBV-specific prophylaxis remains unclear. It is also uncertain whether additional immunoprophylactic measures, such as administering HepBvac and/or HBIG doses beyond conventional schemes, reduce neonatal incidence of OBI.

In the current study, the high-sensitivity nucleic acid amplification assay combined with detection of virus signals by nucleic acid hybridization (NAH) (*i.e.,* PCR/NAH) with sensitivity below 10 HBV genome copies per one mL of serum or plasma or per one microgram of total DNA [5,21,22] was applied to assess OBI incidence in infants who received enhanced passive-active HBV immunoprophylaxis soon after birth to HBsAg-positive mothers treated or not with DAA before parturition. Among the results obtained, the most prominent was the finding that the majority (84.6%) of the children investigated remained HBV reactive during the 24-month follow-up after birth, indicating that the prophylaxis used was only marginally effective in preventing OBI. In addition, sequencing of the HBV preS-S genomic fragments from the blood of children and maternal peripheral and umbilical cord blood revealed HBV sequence homology between infants and umbilical cord blood mononuclear cells but divergence from maternal HBV sequences in 4 of the 5 mother-child pairs examined. This may suggest that some HBV variants in newborns with OBI might be of fetal and/or placental origin.

## Participants and Methods

### Mother-Infant Pairs

Thirteen pregnant women chronically infected with HBV who attended the Antenatal Clinic at the Tianjin Second People’s Hospital (Tianjin) between October 2014 to July 2016 and their infants were investigated. The following groups of mothers were excluded: (1) less than 18 years old and older than 45 years; (2) drinking more than 20 g of pure ethanol per day; (3) co-infected with another virus or with co-existing another infectious disease, (4) having drug-induced liver injury; (5) with autoimmune hepatitis; (6) with decompensated liver disease, and (7) with liver cancer of any kind. The mothers were divided into three study groups.

Group A (n = 4) included mothers whose serum HBV DNA load during pregnancy was consistently lower than 500 HBV genome copies, also called virus genome equivalents (vge), per mL and who did not receive anti-HBV DAA therapy. Mothers in Group B (n = 8) had serum HBV DNA levels initially greater than 1.5 x 10^6^ vge/mL and were treated with telbivudine (LdT; 600 mg qd) from 24-28 weeks of pregnancy that was discontinued postpartum. In this group, HBV load declined to below 500 vge/mL in 4 mothers, as measured at approximately one month before parturition, while continued to be above 1 x 10^3^ vge/mL in four others (Table 1). A mother with HBV DNA loads repeatedly exceeded 1 x 10^6^ copies/mL and carrying 1.5 x 10^8^ vge/mL one month before parturition, who had refused anti-HBV therapy, was included in Group C (Table 1). All women were serum HBsAg positive throughout pregnancy and up to delivery.

**Table 1.**
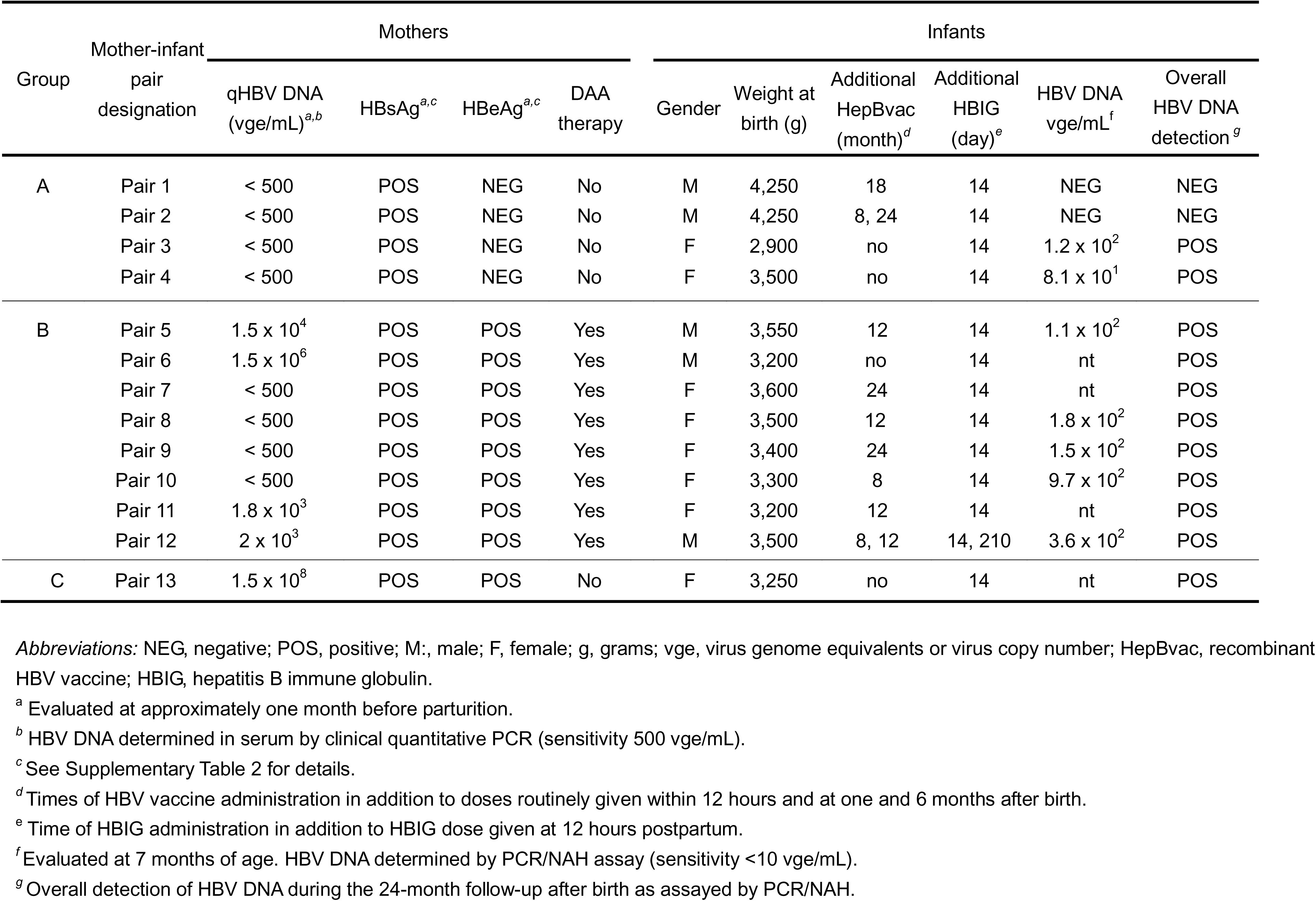
Selected markers of HBV infection in 13 mothers with chronic HBV infection treated or not with DAA prior to delivery and HBV-specific immunoprophylaxis and HBV DNA detection in their children.

| Group | Mother-infant pair designation | Mothers |  |  |  | Infants |  |  |  |  |  |
| --- | --- | --- | --- | --- | --- | --- | --- | --- | --- | --- | --- |
|  |  | qHBV DNA (vge/mL)<sup>a,b</sup> | HBsAg<sup>a,c</sup> | HBeAg<sup>a,c</sup> | DAA therapy | Gender | Weight at birth (g) | Additional HepBvac (month)<sup>d</sup> | Additional HBIG (day)<sup>e</sup> | HBV DNA vge/mL<sup>f</sup> | Overall HBV DNA detection<sup>g</sup> |
| A | Pair 1 | < 500 | POS | NEG | No | M | 4,250 | 18 | 14 | NEG | NEG |
|  | Pair 2 | < 500 | POS | NEG | No | M | 4,250 | 8, 24 | 14 | NEG | NEG |
|  | Pair 3 | < 500 | POS | NEG | No | F | 2,900 | no | 14 | 1.2 x 10<sup>2</sup> | POS |
|  | Pair 4 | < 500 | POS | NEG | No | F | 3,500 | no | 14 | 8.1 x 10<sup>1</sup> | POS |
| B | Pair 5 | 1.5 x 10<sup>4</sup> | POS | POS | Yes | M | 3,550 | 12 | 14 | 1.1 x 10<sup>2</sup> | POS |
|  | Pair 6 | 1.5 x 10<sup>6</sup> | POS | POS | Yes | M | 3,200 | no | 14 | nt | POS |
|  | Pair 7 | < 500 | POS | POS | Yes | F | 3,600 | 24 | 14 | nt | POS |
|  | Pair 8 | < 500 | POS | POS | Yes | F | 3,500 | 12 | 14 | 1.8 x 10<sup>2</sup> | POS |
|  | Pair 9 | < 500 | POS | POS | Yes | F | 3,400 | 24 | 14 | 1.5 x 10<sup>2</sup> | POS |
|  | Pair 10 | < 500 | POS | POS | Yes | F | 3,300 | 8 | 14 | 9.7 x 10<sup>2</sup> | POS |
|  | Pair 11 | 1.8 x 10<sup>3</sup> | POS | POS | Yes | F | 3,200 | 12 | 14 | nt | POS |
|  | Pair 12 | 2 x 10<sup>3</sup> | POS | POS | Yes | M | 3,500 | 8, 12 | 14, 210 | 3.6 x 10<sup>2</sup> | POS |
| C | Pair 13 | 1.5 x 10<sup>8</sup> | POS | POS | No | F | 3,250 | no | 14 | nt | POS |

*Abbreviations:* NEG, negative; POS, positive; M:, male; F, female; g, grams; vge, virus genome equivalents or virus copy number; HepBvac, recombinant HBV vaccine; HBIG, hepatitis B immune globulin.
<sup>a</sup> Evaluated at approximately one month before parturition.
<sup>b</sup> HBV DNA determined in serum by clinical quantitative PCR (sensitivity 500 vge/mL).
<sup>c</sup> See Supplementary Table 2 for details.
<sup>d</sup> Times of HBV vaccine administration in addition to doses routinely given within 12 hours and at one and 6 months after birth.
<sup>e</sup> Time of HBIG administration in addition to HBIG dose given at 12 hours postpartum.
<sup>f</sup> Evaluated at 7 months of age. HBV DNA determined by PCR/NAH assay (sensitivity <10 vge/mL).
<sup>g</sup> Overall detection of HBV DNA during the 24-month follow-up after birth as assayed by PCR/NAH.

### Perinatal Hepatitis B Immunoprophylaxis

All infants received 100 IU of HBIG (Chengdu Rongcheng Pharmaceuticals, Chengdu, PR China) intramuscularly *(i.m.)* within 12 hours after birth. An additional dose of 100 IU was administered 14 days after birth or as indicated (Table 1). As well, recombinant HepBvac (GlaxoSmithKline Biologicals SA, Rixensart, Belgium) was given *i.m.* at 10 mg (0.5 mL) per injection within the first 12 hours postpartum, and one and 6 months thereafter. If antibodies to HBsAg (anti-HBs) declined below 200 mIU/mL, the vaccine was given at 8 months of age and/or at later time points (Table 1).

### Samples Collection

For this study, blood samples were obtained from mothers’ peripheral venous blood approximately one month prior to parturition. Maternal peripheral blood plasma (MPBP) and maternal peripheral blood mononuclear cells (MPBC; otherwise commonly called peripheral blood mononuclear cells or PBMC), as well as umbilical cord blood plasma (UCBP) and umbilical cord blood mononuclear cells (UCBC) were collected at delivery. MPBC and UCBC were isolated from acid-citrate-dextrose (ACD)-treated blood applying a Ficoll-HyPague density gradient centrifugation, as reported [5,23], and their pellets stored at −80 ℃ until analysis. Blood samples from infants were obtained at 2 days of life and at 2, 7, 12 and 24 months of age. It should be noted that it was not always feasible to secure samples at all times indicated due to family unwillingness to subject children to needle punctures or issues with children’s transportation to hospital. Another constraint was the limited availability of sample material from children for HBV DNA detection by PCR/NAH since testing by clinical assays was accepted to be a priority. Considering blood samples from children, which were collected in the absence of anticoagulant, sera accumulating above the blood clots and the clots containing both blood cells and serum leftovers, called as whole blood clot (WBC), were collected separately and stored at −80 ℃. However, for the high-sensitivity HBV detection by PCR/N in infants AH, WBC were used only.

The complete list of abbreviations identifying the samples investigated is as follows: MPBP or MP, maternal peripheral blood plasma or maternal plasma; MPBC or MC, maternal peripheral blood mononuclear cells or maternal cels; UCBP, umbilical cord blood plasma; UCBC or UC, umbilical cord blood mononuclear cells or umbilical cells; WBC02 or B02, whole blood clot from infant at 2 days of life; WBC2 or B2, whole blood clot from 2 months old infant; WBC7 or B7, whole blood clot from 7 months old infant; WBC12 or B12, whole blood clot from 12 months old infant, and WBC24 or B24, whole blood clot from 24 months old infant.

### Evaluation of Immunovirological Markers of HBV Infection

HBV infection markers in circulating blood were evaluated using the Roche Diagnostics GmbH system (Mannheim, Germany) and Cobas E601 electrochemiluminescence analyzer (Basel, Switzerland). Following the manufacturer’s instructions, HBsAg was considered negative when reading was < 0.05 IU/mL, anti-HBs negative when < 10 mIU/mL, HBV e antigen (HBeAg) negative when cut-off index (COI) was < 1.0, antibodies to HBeAg (anti-HBe) considered negative when COI ≥ 1.0, and anti-HBc negative when COI was > 1.0.

### Liver Biochemical Evaluations

Evaluation of liver biochemical function in mothers included testing for serum alanine aminotransferase (ALT; normal range 7 - 40 U/L), aspartate aminotransferase (AST; 13 - 35 U/L), gamma-glutamyl transpeptidase (GGT; 7 - 45 U/L), alkaline phosphatase (ALP, 50 - 135 U/L), total bilirubin (TBIL; 3.4 - 17.1 µmol/L), direct bilirubin (DBIL; 0 – 6.84 µmol/L), total protein (TP; 62 85 g/L), albumin (ALB; 35 - 55 g/L), and albumin/globulin ratio (A/G; 1.2 - 2.5). All the assays were performed with Hitachi 7180 automatic biochemical analyzer (Tokyo, Japan).

### HBV DNA Testing by Clinical Quantitative PCR Assay

Total DNA was extracted from 200 µL serum or plasma of mothers and of their infants using the High Pure Viral Nuclei Acid kit (Roche Diagnostics, Branchburg, NJ, USA). HBV DNA load was determined by quantitative PCR with HBV surface (S) gene primers using a LightCycler Roche LC480 instrument (Roche Diagnostics, Penzberg, Germany). The assay sensitivity was 500 vge/mL. All samples from mothers and children were evaluated using this test except samples collected from children at the age of 2 years which were tested at the assay sensitivity of 100 vge/mL.

### HBV DNA Detection by High-Sensitivity PCR/NAH Assay

DNA was extracted from mothers’ MPBC and UCBMC and from infants’ WBC using a QIAamp DNA Blood Kit (Qiagen GmbH, Hilden, Germany) and from one mL of serum or plasma obtained from peripheral and cord blood of mothers. 2.5-3 μg DNA from MPBC, UCBMC and WBC, and DNA recovered from the whole volume of serum or plasma served as templates for direct PCR amplification and then 10 µL of the product was used for nested PCR. HBV S and X gene-specific primers were used in both runs under conditions reported previously [5,21,22] (Supplementary Table 1). Each amplification reaction was supplemented with positive amplification and negative contamination controls, and with ten-fold dilutions of complete recombinant HBV as quantitation standards, as reported [5,21]. To verify specificity of the amplified HBV signals and to augment the assay sensitivity, NAH (*i.e.,* Southern blot hybridization analysis) was used [5,21,22]. The sensitivity of nested PCR coupled with NAH (*i.e.,* PCR/NAH) was 5 - 10 vge per mL of serum or plasma or per one microgram of total DNA. This sensitivity was closely comparable to that of similar assays designed to detect low levels of viral genomes in other studies [23–28].

### HBV Sequencing and Phylogenetic Analysis

Either direct or clonal sequence analysis of the HBV preS-S region encompassing the 360-bp fragment spanning between nucleotides 3124-364, as enumerated based on the HBV sequence with GenBank accession number LC057377, was done to determine HBV sequence variations between mothers, mononuclear cells from umbilical cord blood and their offspring. The amplicons were purified and directly sequenced or cloned using the TOPO-TA cloning system (Invitrogen, Carlsbad, CA, USA) and then sequenced, as described before [27–29]. Thus, direct sequencing was done of PCR products obtained from 9/F and 12/F mothers, their UCBC, and WBC samples from their children. Regarding clonal sequencing, up to 11 randomly selected clones derived from nPCR products obtained from 8/F, 10/F and 11/F mothers, UCBC, and WBC of their children who were followed for up to 12 months after birth were analyzed. The clones were sequenced in both directions using universal forward and reverse M13 primers by applying an Ion S5 Next-Generation Sequencing System (Termo Fisher Scientific, Waltham, MA, USA). Analysis of the sequencing data was done with help of the CLC Genomic Workbench software (Qiagen). Neighbor-joining tree phylogenetic construction was based on the Tamura-Nei model and completed using the MEGAv7 software (accessed at www.magasoftware.net).

### Statistical Analysis

Statistical analyses were done using the Statistical Package for Social Science (SPSS) for Windows, Version 21.0 (SPSS Inc., Chicago, IL, USA). Non-normal variables were expressed as median with minimum and maximum values and analyzed using Mann-*t* test. Candidate variables with a *P* value < 0.05 on univariate analysis were included in the multivariate logistic regression model. All statistical tests were two-tailed. Values of *P* < 0.05 were considered as statistically significant.

## Results

### HBV Infection and Liver Function in Pregnant Mothers

Serological markers of HBV infection and serum HBV DNA loads were examined in mothers approximately one month prior to delivery. Testing for HBsAg confirmed that all (n = 13) were antigen-positive at that time point (Table 1 and Supplementary Table 2). One mother (7/F) also was reactive for anti-HBs (Supplementary Table 2). Nine were serum HBeAg-positive, although 8 received LdT therapy (Group B; Table 1), while the remaining 4 DAA-untreated antigen-negative (Group A; Table 1). Three 3 of those 4 were anti-HBe-positive (Supplementary Table 2). Eleven of 13 mothers tested for anti-HBc were antibody reactive (Supplementary Table 2). At one month before parturition, HBV was detected at levels above 1 x 10^3^ vge/mL in 4 of 8 mothers treated with LdT and in a mother belonging to Group C (Table 1). The remaining 8 woman were HBV DNA-negative when tested by the clinical assay with the sensitivity of 500 vge/mL (Table 1 and Supplementary Table 2). However, 7 of them carried HBV at the time of parturition in MPBP and/or MPBC and in UCBC at levels below 1.9 x 10^2^ vge/mL when examined by the high-sensitivity HBV DNA detection assay (see below).

Biochemical indicators of liver function were within normal ranges in all 13 mothers at a month prior to delivery with the exception of alkaline phosphatase (ALP) levels in 3 mothers (Supplementary Table 2). The level of ALP above normal limit of 135 U/L could be due to mothers’ fat-rich diet and/or the enzyme release during fetal bone development [30]. Otherwise, there were no statistically significant differences in enzymatic indicators of liver function between HBeAg-positive (n = 9) and HBeAg-negative (n = 4) mothers or between mothers with the higher (*i.e.,* > 500 vge/mL) (n = 5) and the lower (*i.e.,* < 500 vge/mL) (n = 8) HBV loads. However, when alanine aminotransferase (ALT) levels in mothers with the higher HBV loads (*i.e.,* ALT median 27.0 U/L with inter-quartile range of 21.0 - 35.5 U/L; n = 5) were compared to those in mothers with the lower HBV loads (*i.e.,* ALT median 18.0 U/L with inter-quartile range of 15.2 - 19.0 IU; n = 8) a statistically significant difference became evident (*P* = 0.039). This may indicate that a very mild liver necroinflammation endured in mothers with the higher HBV loads and implies that ALT above 18 U/L might be considered as elevated in HBV-infected pregnant females. This collaborates with the notion that the upper normal ALT level for healthy females should be 18 U/L [31].

### HBV Detection in Maternal Plasma and Cells from Peripheral and Umbilical Cord Blood

We had a rare opportunity to compare HBV detection in maternal peripheral and umbilical cord blood plasma (*i.e.,* MPBP and UCBP) and mononuclear cells isolated from these two compartments (*i.e.,* MPBC and UCBC) at the time of parturition and analyzed them for HBV presence by the high sensitivity PCR/NAH assay. The overall rate of HBV detection in MPBP samples was 50% (6/12), and it was 25% (1/4) for mothers belonging to Group A (*i.e.,* LdT-untreated) and 57.1% (4/7) for those from Group B (*i.e.,* LdT-treated). For MPBC, the overall rate of HBV detection was 84.6% (11/13) and for mothers from Group A was 50% (2/4), while for those from Group B 100% (8/8). MPBP and MPBC from LdT-untreated 13/F mother (Group) were both HBV DNA-positive. Further, the overall rate of HBV detection in UCBC was the same as in MPBC, *i.e.,* 84.6% (11/13), and the rates for Group A and Group B were 50% (2/4) and 100% (8/8), respectively. Curiously, all UCBP samples were found to be HBV DNA-negative. This was confirmed by repeated testing with primers specific to HBV S (surface), X, and C (core) genes (Supplementary Table 1). A reason for this is unknown, but it could be related to the presence in umbilical plasma of a factor co-isolating with DNA that inhibits PCR, as it was shown for several substances [32,33]. Thus, maternal mononuclear cells from either peripheral or umbilical cord blood gave the greatest and identical rates of HBV detection (∼85%), the rate was lower in maternal peripheral plasma (50%), while umbilical cord plasma was unsuitable for HBV DNA identification in this study. These data confirmed that circulating mononuclear cells, which include T cells, B cells, NK cells and monocytes [34], are the most reliable material for detection of low levels of HBV. This is consistent with findings in the model HBV infection in woodchucks, hepatitis C virus (HCV) infection and in a number of other persistent infections with lymphotropic viruses [3,35–38].

### Overall Characteristics of HBV Infection Markers in Infants Receiving Immunoprophylaxis

All children were serum HBsAg negative at 2 days and at 2 and 7 months (n =13), 12 months (n =12), and 24 months (n = 11) after birth (Table 2 and Supplementary Table 3). The rates of anti-HBs positivity were 15.4% (2/13) at 2 days postpartum, 100% (13/13) at 2 months of age, 92.3% (12/13) at 7 months, 100% (12/12) at 12 months, and 100% (11/11) at 24 months (Table 2). Two newborns at 2 days of life carried anti-HBs but at levels below 100 mIU/mL (Supplementary Table 3). HBeAg was detected in 53.8% (7/13) at 2 days and 15.4% (2/13) at 2 months, but was not found during the remaining 24-month observation period (Table 2). Anti-HBe reactivity was present in 46.1% (6/13) at 2 days after birth, 30.8% (4/13) at 2 months, and 23.1% (3/13) at 7 months, but was not detected at 12 and 24 months of age (Table 2). Anti-HBc were positive in all 13 neonates 2 days and 2 months.

**Table 2.** Overall profiles of HBV infection markers in infants during the 24-month follow-up after birth to HBsAg-positive mothers.

| HBV infection marker | Time of follow-up |  |  |  |  |
| --- | --- | --- | --- | --- | --- |
|  | 2 days | 2 months | 7 months | 12 months | 24 months |
| HBsAg<sup>a</sup> | 0/13 (0%) | 0/13 (0%) | 0/13 (0%) | 0/12 (0%) | 0/11 (0%) |
| Anti-HBs<sup>a</sup> | 2/13 (15.4%) | 13/13 (100%) | 12/13 (92.3%) | 12/12 (100%) | 11/11 (100%) |
| HBeAg<sup>a</sup> | 7/13 (53.8%) | 2/13 (15.4%) | 0/13 (0%) | 0/12 (0%) | 0/11 (0%) |
| Anti-HBe<sup>a</sup> | 6/13 (46.1%) | 4/13 (30.7%) | 3/13 (23.1%) | 0/12 (0%) | 0/11 (0%) |
| Anti-HBc<sup>a</sup> | 13/13 (100%) | 13/13 (100%) | 11/13 (84.6%) | 7/12 (58.3%) | 10/11 (90.9%) |
| HBV DNA<sup>b</sup> | nt | 5/7 (71.4%) | 7/9 (77/8%) | 6/10 (60%) | 2/3 (66.7%) |

*Abbreviations:* nt, not tested.
<sup>a</sup> Evaluated by Cobas E601 electrochemiluminescence immunoassays (Roche Diagnostics, Basel, Switzerland).
<sup>b</sup> Evaluated by PCR/NAH assay (sensitivity <10 vge/mL).

Then, their detection declined to 84.6% (11/13) at 7 months and further to 58.3% (7/12) at 12 months, and rebounded to 90.9% (10/11) at 24 months of age (Table 2). As indicated, HBV DNA was not detected throughout the 24-month follow-up by the available clinical laboratory assay, but it was detected when the high sensitivity PCR/NAH was applied (Table 2). Thus, HBV was identified in 71.4% (5/7) at 2 months, 77.8% (7/9) at 7 months, and in 60% (6/10) at 12 months of age. At 24 months, samples from 3 children were available for testing and 2 of them were HBV DNA positive (Table 2). The estimated WHV DNA loads during the two-year follow-up ranged from 8.1 x 10^1^ to 2.1 x 10^3^ vge/mL (median 1.35 x 10^2^ vge/mL with inter-quartile range of 1.1 x 10^2^ - 2.07 x 10^2^ vge/mL) (Supplementary Table 3). Only two (15.4%; 2/13) infants, both born to LdT-untreated, HBeAg-negative mothers, were HBV DNA negative throughout 2-year observation after birth (Table 2). The remaining eleven (84.6%; 11/13) were HBV genome reactive at one or multiple time points during this period. The highest rate of HBV detection of 77.8% (7/9) was at the age of 7 months (Table 2). Considering the total number of WBC samples examined, 68.9% (20/29) were found HBV DNA positive by the high sensitivity PCR/NAH assay.

### Profiles of HBV Infection Markers in Infants Born to Mothers with Low or High HBV Load before Parturition

To recognize the potential impact of maternal infection on the serological profiles of HBV infection markers in children, the presence of individual HBV markers was reanalyzed taking into consideration maternal HBV load detected at about one month prior to delivery. In this regard, children were grouped based on maternal HBV load independently of whether or not mothers were treated with LdT (Table 1). The group with the maternal HBV loads below 500 vge/mL was designated as the maternal low virus load group or the MLVL group and included infants: 1/M, 2M, 3/F, 4/F, 7/M, 8/F, 9/F, and 10/F (n = 8). The group with the maternal viral loads greater than 500 vge/mL was designated as the maternal high virus load group or the MHVL group and included infants: 5/M, 6/F, 11/F, 12/M, and 13/M (n = 5) (Table 1 and Supplementary Table 3).

As summarized in Table 3 and presented in detail in Supplementary Table 3, anti-HBs were detected at 2 days postpartum at levels below 100 mIU/mL in one newborn in each group. This corresponded to rates of 12.5% (1/8) and 20% (1/5) for the MLVL and MHVL groups, respectively. From 2 months onwards, anti-HBs occurred at levels higher than 200 mIU/mL in almost all infants. The exception was 2/M from the MLVL group in whom anti-HBs were below 200 mUL/mL at 2 months and negative at 7 months of age, although HBIG was given within 12 hours and at 2 weeks after birth and two additional doses of HepBvac were administered at 8 and 24 months of age (Table 1). At 7 months, anti-HBs also dropped below 100 mIU/mL in 10/F from the MLVL group and in 12/M from the MHVL group compared to the levels of more than 300 mIU/mL at 2 months (Supplementary Table 3). This prompted injections of additional doses of HepBvac to both infants and 200 IU of HBIG to 12/M (Table 1). These interventions did not result in any substantial increase in the anti-HBs levels, which were slightly above 100 mIU/mL at 12 months in 10/F and remained below 100 mIU/mL at 12 and 24 months in 12/M (Supplementary Table 3). In addition, 7/M and 9/F from the MLVL group, and 11/F from the MHVL group were re-injected with HepBvac at 24 or 12 months, respectively, due to decreases in anti-HBs. Some infants with anti-HBs declining below 200 IU were not re-vaccinated due to temporal health issues, such as elevated body temperature or diarrhea.

**Table 3.** Profiles of individual HBV infection markers over the 24-month follow-up of infants born to mothers with low or high HBV loads prior to parturition.

| HBV infection marker | Mother subgroup | Time of follow-up |  |  |  |  |
| --- | --- | --- | --- | --- | --- | --- |
|  |  | 2 days | 2 months | 7 months | 12 months | 24 months |
| Anti-HBs<sup>a</sup> | MLVL | 1/8 (12.5%) | 8/8 (100%) | 7/8 (87.5%) | 7/7 (100%) | 6/6 (100%) |
|  | MHVL | 1/5 (20%) | 5/5 (100%) | 5/5 (100%) | 5/5 (100%) | 5/5 (100%) |
| HBeAg<sup>a</sup> | MLVL | 3/8 (37.5%) | 0/8 (0%) | 0/8 (0%) | 0/7 (0%) | 0/6 (0%) |
|  | MHVL | 4/5 (80%) | 2/5 (40%) | 0/5 (0%) | 0/5 (0%) | 0/5 (0%) |
| Anti-HBe<sup>a</sup> | MLVL | 5/8 (62.5%) | 4/8 (50%) | 3/8 (37.5%) | 0/7 (0%) | 0/6 (0%) |
|  | MHVL | 1/5 (20%) | 0/5 (0%) | 0/5 (0%) | 0/5 (0%) | 0/5 (0%) |
| Anti-HBc<sup>a</sup> | MLVL | 8/8 (100%) | 8/8 (100%) | 6/8 (75%) | 5/7 (71.4%) | 6/6 (100%) |
|  | MHVL | 5/5 (100%) | 5/5 (100%) | 5/5 (100%) | 2/5 (40%) | 4/5 (80%) |
| HBV DNA<sup>b</sup> | MLVL | nt | 3/4 (75%) | 5/7 (71.4%) | 4/8 (50%) | 0/1 (0%) |
|  | MHVL | nt | 2/3 (66.6%) | 2/2 (100%) | 2/2 (100%) | 2/2 (100%) |

*Abbreviations:* MLVL, mothers with low HBV load (i.e., < 500 vge/mL) at about one month before parturition; MHVL, mothers with high HBV load (i.e., > 500 vge/mL) at approximately one month before parturition; nt, not tested.
<sup>a</sup> Evaluated by Cobas E601 electrochemiluminescence immunoassay system (Roche Diagnostics, Basel, Switzerland).
<sup>b</sup> Evaluated by in house PCR/NAH assay (sensitivity <10 vge/mL).

Interestingly, the HBeAg reactivity was detected at day 2 in 37.5% (3/8) of newborns from mothers with low HBV load prior to delivery, but in 80% (4/5) of these born to women with higher HBV load, but this difference was not statistically significant (*P* = 0.26) (Table 3 and Supplementary Table 3). Further, HBeAg remained present at the age of 2 months in 40% (2/5) of infants belonging to the MHVL group, but not in those from the MLVL group, and was not detected onward in either group (Table 3). The profile of anti-HBe reactivity was in general inverse to that of HBeAg. Hence, the antibodies were detected at day 2 in 62.5% (5/8) of newborns from mothers with low HBV loads and in just one of five (20%) born to females with higher HBV levels (Table 3 and Supplementary Table 3). While anti-HBe remained detectable at 2 months in 50% (4/8) and at 7 months in 37.5% (3/8) of infants from MLVL group, the antibodies were negative in infants belonging to the MHVL group at any other time of follow-up than 2 days (Table 3).

Anti-HBc circulated in all newborns at 2 days and 2 months after birth (Table 3 and Supplementary Table 3), and they were present in all (5/5) infants belonging to the MHVL group and in 75% (5/8) of those from MLVL at 7 months. At 12 months, anti-HBc positivity rates declined to 71.4% (5/7) in the MLVL children and to 40% (2/5) in the MHVL group. Subsequently, anti-HBc rebounded and were detected at 24 months in all (6/6) infants belonging to the MLVL group and in 80% (4/5) of these born to mothers with higher HBV loads before parturition (Table 3).

As already pointed out, all infants were seemingly negative for HBV DNA during the 2-year follow-up when tested by clinical laboratory test. Testing by PCR/NAH brought overall HBV DNA detection at a higher frequency in samples collected from the MHVL infants than in those from the MLVL group, *i.e.,* 88.9% (8/9) *vs.* 60% (12/20), respectively, but these values were not statistically different (*P* = 0.2) (Table 3 and Supplementary Table 3).

In summary, infants born to mothers with relatively low or high HBV loads prior to parturition showed different profiles of HBV infection markers during the 2-year observation after birth. While HBsAg was uniformly undetectable, the HBeAg and anti-HBe profiles were the most distinct between infants belonging to MLVL or MHVL group. Anti-HBc tended to have biphasic profiles in both groups and their levels decreased between 7 to 12 months after birth and rebound at the end of the observation period. This may suggest a decline in anti-HBc transferred from mothers and initiation of own synthesis of the antibodies due to ongoing HBV infection. HBV DNA was detected in the majority of samples available for analysis by the high sensitivity assay employed. The greatest positivity rate was identified in children around 7 months of age who were born to mothers with the high HBV load prior to delivery.

### Profiles of HBV Infection Markers in Infants born to HBsAg-Positive Mothers Treated or Not with DAA before Parturition

To evaluate the effect of gestational DAA treatment on the HBV infection in newborns, the profiles of immunovirolgical markers of HBV infection and HBV DNA detection were compared in infants born to the LdT-treated and the untreated mothers. Accordingly, the group of infants born to the treated mothers (Group B; Table 1) included 5/M, 6/F, 7/M, 8/F, 9/F, 10/F, 11/F, and 12/M (n = 8) and was designated as the infant-from-treated-mother group or the IFTM group. Infants born to the untreated mothers (Groups A and C; Table 1) constituted the infant-from-untreated-mother group or the IFUTM group and included 1/M, 2M, 3/F, 4/F, and 13/M (n = 5). Since infants born to mothers who reached lower HBV load (*i.e.,* < 500 vge/mL) due to LdT therapy (n = 4) and these born to the LdT-treated mothers in which the treatment was not successful in lowering HBV below 500 vge/mL (n = 4) constituted the IFTM group (Table 3), these two subgroups were also compared.

Considering the anti-HBs profile, the antibodies were detected 2 days after birth in two newborns in the IFTM group (25%; 2/8) of which one belonged to the MLVL subgroup and another to the MHVL group, while no anti-HBs reactivity was found in newborns from the IFUTM group (Table 4). Subsequently, anti-HBs were measurable in all infants, except 2/M at 7 months of age, throughout the remaining 2-year observation period (Supplementary Table 3).

**Table 4.**
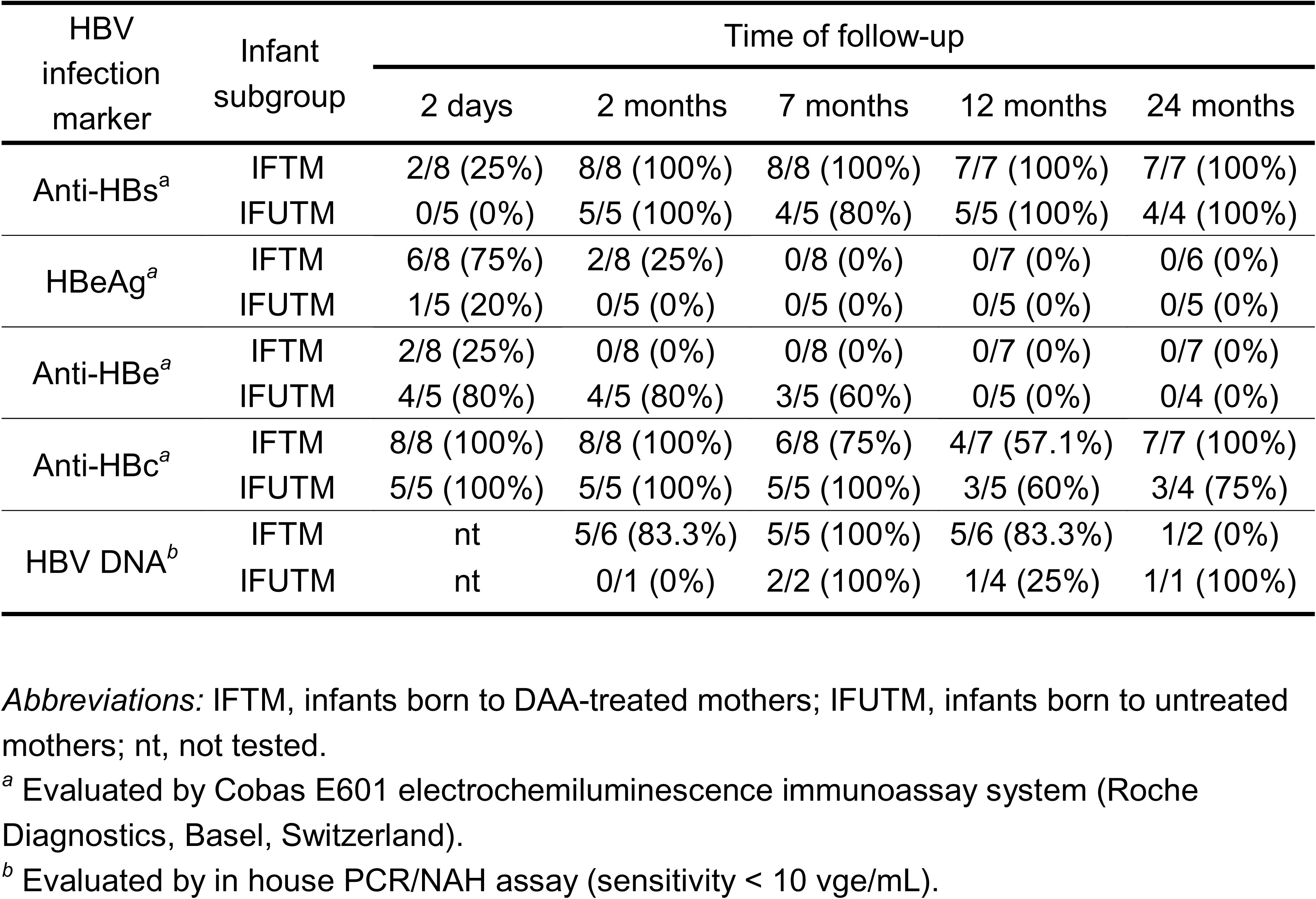
Profiles of individual HBV infection markers over the 24-month follow-up of infants born to mothers treated or not with a DAA compound prior to parturition.

HBeAg was positive on day 2 in 75% (6/8) of newborns in the IFTM group, among whose 3 belonged to MLVL and 3 others to the MHVL subgroup, and in 1/5 (20%) of newborns from the IFUTM group (Table 4 and Supplementary Table 3). The antigen was also detectable at 2 months in 25% (2/8) in the IFTM group and both positive infants were from the MLVL subgroup. After this point, all children became HBeAg-negative until the end of follow-up (Table 4). Anti-HBe showed opposing profiles to the HBeAg detection. Hence, while the antibodies were positive at 2 days of life in 25% (2/8) of newborns in the IFTM group (one of each in MLVH and MHVL subgroups) and in 80% (4/5) of those in the IFUTM group, they were not detected at any later time points in the IFTM group. In contrast, anti-HBe remained detectable for up to 7 months in 3/5 (60%) in infants belonging to the IFUTM group (Table 4 and Supplementary Table 3).

Anti-HBc were identified in all newborns at 2 days and 2 months of age from the IFTM (8/8) and IFUTM (5/5) groups (Table 4). At 7 months, 75% (6/8) in the IFTM group, among them 4 belonged to the MHVL subgroup and 2 to the MLVL subgroup, and all (5/5) in the IFUTM group were anti-HBc reactive. Anti-HBc reached the lowest positivity rate at 12 months after birth in both IFTM (57.1%; 4/7) and IFUTM (60%; 3/5) groups. The antibody rates rebounded 24 months to 100% (7/7) and 75% (3/4), respectively (Table 4).

Regarding HBV DNA testing by PCR/NAH, 19 samples from the IFTM group and 8 from the IFUTM group collected during the 24-month follow-up were available for examination, and 16 (84.2%) and 8 (50%) of them were identified as positive, respectively. Among the 16 positive samples from infants in the IFTM group, 56.2% (9/16) belonged to the MLVL subgroup and 43.7% (7/16) to the MHVL subgroup (*P* = 0.72). Considering the rate of HBV carriage among infants at 12 and/or 24 months of age, 2 samples from the 2 infants tested who belonged to the IFUTM group and 6 from the 6 tested from the IFTM group were HBV DNA positive. Among the 6 infants from the IFTM group, 3 were from the MLVL subgroup and 3 others from the MHVL subgroup (Supplementary Table 3). Taken together, the data indicated that LdT anti-HBV treatment mothers did not influence OBI incidence in their children.

Considering overall profiles of HBV infection markers, the HBeAg and anti-HBe patterns were most divergent over time between infants belonging to the IFTM or IFUTM groups. While the HBeAg and anti-HBe profiles in the IFTM group more closely corresponded to the those in the MHVL group, the profiles in the IFUTM group more resembled those in the MLVL group (Tables 3 and 4). It could also be noted that: (1) both IFUTM and MLVL groups were characterized by a very brief period of HBeAg positivity and the presence of anti-HBe ceased after 7 months of age; (2) both IFTM and MHLV groups displayed HBeAg up to 2 months in about a half of the infants, while anti-HBe vanished shortly after birth and became undetectable from 2 months onwards, and (3) The data suggested that HBeAg and anti-HBe in children were passively transmitted from mothers and neither HBeAg re-appeared nor anti-HBe were *de novo* produced in response to ongoing occult infection during the first 2 years of life.

### Comparison of HBV PreS-S Genomic Sequences from Mothers, Umbilical Cord Blood Cells, and Infants with Occult Infection

In an effort to recognize potential differences in the HBV genomic sequences between mothers and their newborn children, bidirectionally sequenced clones or directly sequenced amplicons of the 360-bp HBV preS-S genomic fragments derived from maternal plasma (*i.e.,* MP) or maternal peripheral blood mononuclear cells (*i.e.,* MC), umbilical cord blood mononuclear cells (*i.e.,* UC), and whole blood of infants between 2 and 12 months of age (*i.e.,* B2, B7 and B12) were comparatively analyzed. In this regard, the HBV sequences identified in infants and UC were first compared to those found in mothers and then the sequences detected in children to those from UC.

Clonal sequencing of the preS-S amplicons from the mother-infant pair referred to as Pair 8 showed essentially identical virus sequence in MP and MC compatible with HBV subgenotype C2 (GenBank AF539983 used as reference) (Figure 1A), with the exception of two mismatched point mutations in MP and one in MC (data not shown). However, the great majority of clones (18/25; 72%) derived from the child at 2, 7 and 12 months of age contained HBV C/D subgenotype (GenBank AY817515 used as reference sequence) (Figure 1A) and displayed multiple (n = 31) point mutations when compared to the maternal virus (data not shown). These mutations represented 8.6% of the 360-bp sequence analyzed. In addition, HBV sequences homologous with the Ce subgenotype (GenBank DQ089793 used as reference) was detected in 2/8 and 5/9 clones (overall 7/25; 28%) derived from the infant at 2 and 12 months of age, respectively, but not in any of the clones from a sample collected at the age of 7 months (Figure 1A). UC sample from Pair 8 was not available for analysis. In general, phylogenetic analysis indicated separate clustering of the HBV sequences in MP and MC from those identified the child samples (Figure 1A).

**Figure 1.**
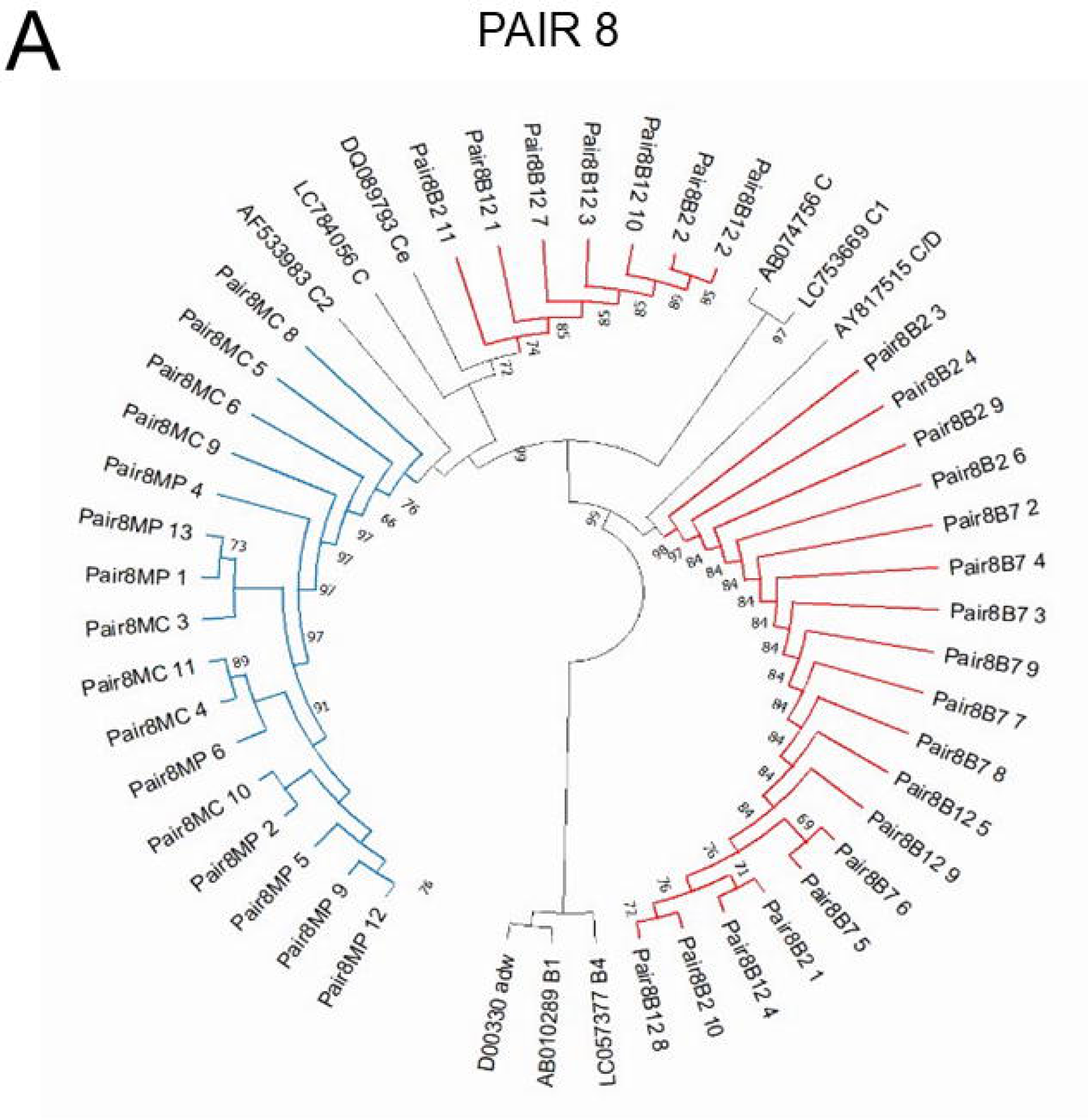
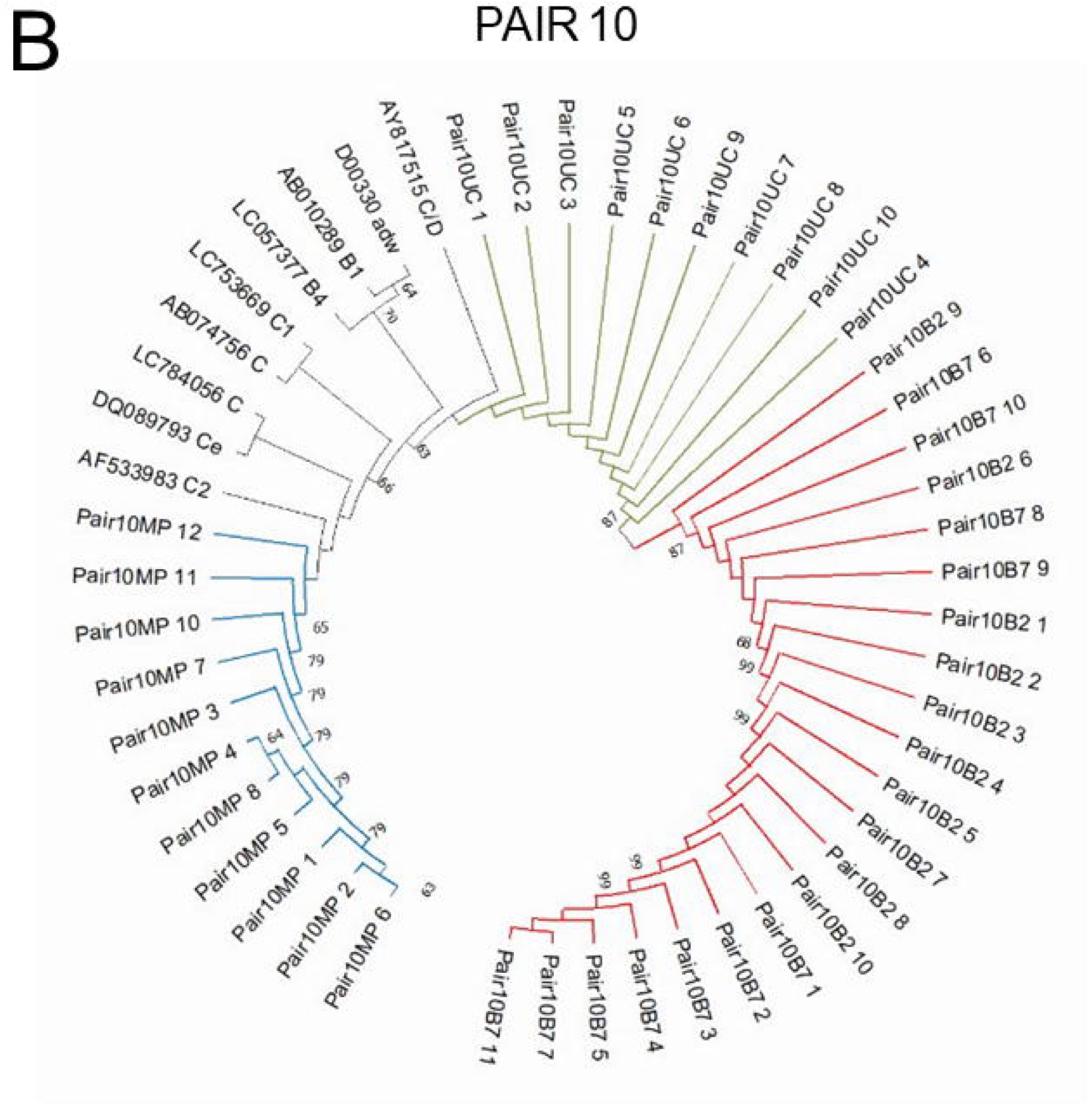
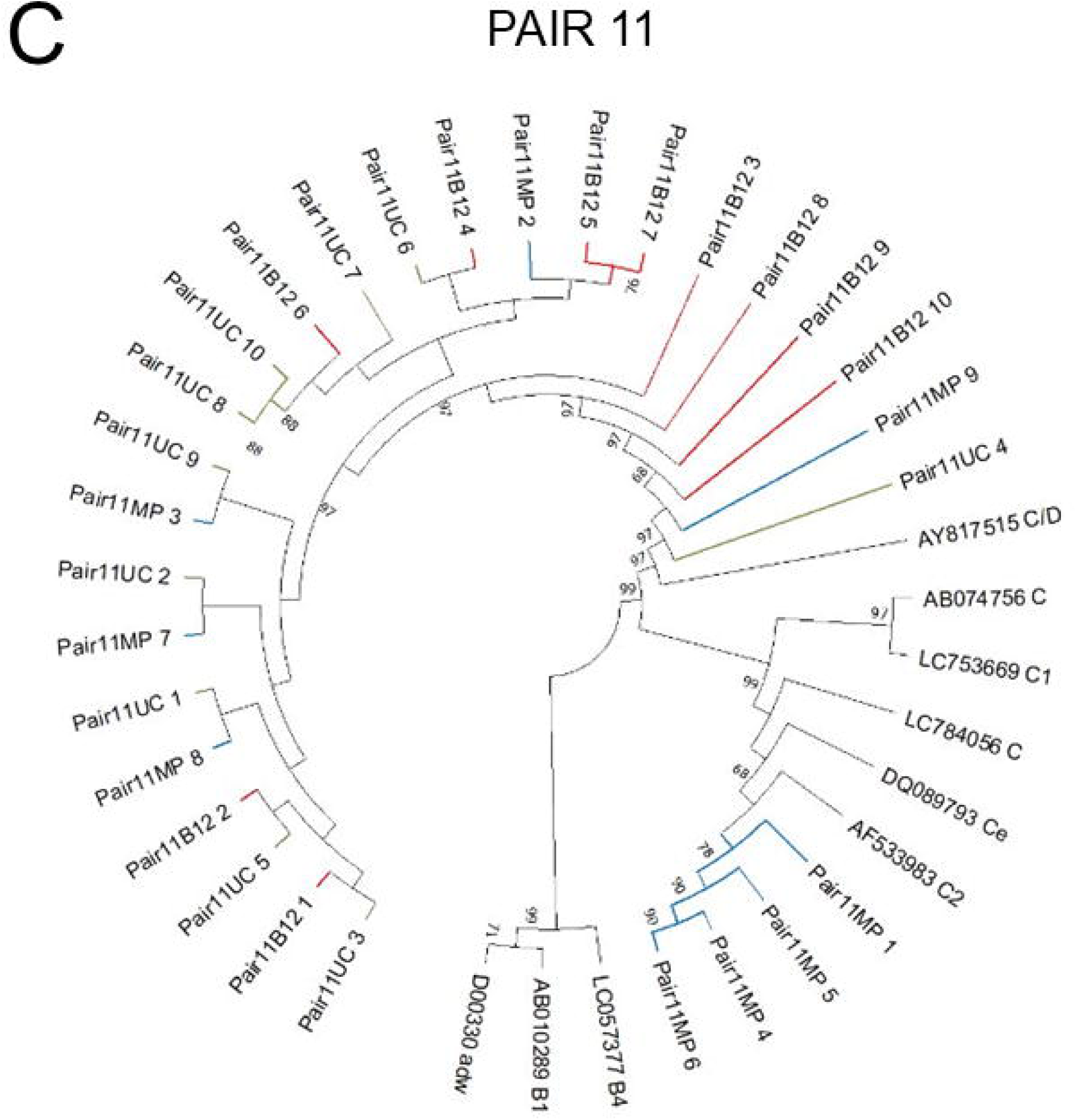
Neighbor-joining phylogenic tree of the cloned 360-bp fragments of the HBV preS-S genomic region derived from samples obtained from mother-child pair 8 (A), pair 10 (B), and pair 11 (C). Samples identification: MP, mother plasma; MC, mother peripheral blood mononuclear cells; UC, umbilical cord blood mononuclear cells; B2, whole blood clot (WBC) containing blood cells and plasma from 2-month-old; B7, WBC from 7-month-old; and B12, from 12-month-old infant. Between 9 and 11 clones from individual samples are shown. Blue lines mark clones from MP and MC, green from UC, red from B2, B7 and B12, and black mark position of equivalent in the size and location nucleotide fragments from 9 GenBank sequences representing different HBV subgenotypes.

MP, UC, B2 and B7 samples were available for analysis from Pair 10 and 10 to 11 clones per sample were sequenced and compared (Figure 1B). The data indicated that the 360-bp preS-S sequences from the child at 2 and 7 months of age and from UC were identical, but they diverged from those occurring in MP. This was a very compelling finding since all 10 of the 10 clones from UC and all clones from B2 and B7 samples displayed the same sequence that was consistent with HBV C/D subgenotype (Figure 1B). This variant sequence diverged at 28 (7.8%) nucleotide positions from the 360-bp fragment identified in all 11 clones originating from MP, which carried C2 subgenotype (Figure 1B).

Likewise, MP, UC and B12 samples from Pair 11 were analyzed. Interestingly, MP displayed two HBV subgenotypes C2 and C/D (Figure 1C). These subgenotypes were found in 4/9 and 5/9 clones from MP analyzed, respectively, and they differed at 27 (7.5%) nucleotides from each other (Figure 1C). Clones from the infant obtained at 12 months of age and from umbilical cord blood mononuclear cells showed the same sequence in all 10 clones examined that was identical with the maternal C/D subgenotype sequence (not shown).

Direct sequencing of the preS-S amplicons was only feasible from the mother-infant pairs designated as Pair 9 and Pair 12. The 360-bp fragments from MC, UC and B12 samples of Pair 9 and from MC, UC and B7 samples of Pair 12 were sequenced and compared to the HBV genotype C reference sequences, and to each other within each pair (data bot shown). Considering Pair 9, maternal mononuclear cells displayed HBV subgenotype C2. There were 21 single nucleotide mutations which were identical in UC and the infant B12 sample but were distinct from the HBV sequence in MC (data not shown). These point mutations constituted 5.8% of the 360-bp preS-S sequence present in MC. Sequences in UC and B12 samples were most closely matched the equivalent C/D subgenotype sequence. Overall, the HBV sequence identified in the Pair 9 infant was identical with that in UC, similarly as it was already uncovered for children belonging to Pair 10 and Pair 11. Regarding Pair 12, analysis of the preS-S fragments from MC and UC and the infant B7 sample showed identical HBV genotype C sequence in all three compartments (data nor shown).

The above data demonstrated the uniformity of a variant mutation within the HBV preS-S sequences in children and the umbilical cord blood mononuclear cells which had clearly diverged from the maternal HBV preS-S sequence in 3 of 5 mother-infant pairs analyzed. In addition, the preS-S sequence in the Pair 8 child was identical with these found in children from Pairs 9, 10 and 11 (Table 5). Thus, among a total 32-point mutations in the preS-S sequence in these children which differentiated from those in their mothers, 21 (65.5%) were shared by all four mother-child pairs. Four other single nucleotide point mutations were common for Pairs 8, 10 and 11 but not Pair 9 and two others for Pairs 8 and 10 but not occurred in Pairs 9 and 11 (Table 5). There were only 5/32 (15.6%) point mutations that were unique for the particular mother-infant pairs and they were found in Pairs 8, 10 and 11. Overall, the results from 4 randomly selected pairs showed surprisingly high degree of HBV variant sequence homology between children and immune cells from umbilical cord blood and, at the same time, a divergence from the equivalent maternal sequences. This may suggest a common mechanism of creation of this virus sequence dichotomy, possibly indicative of the fetal and/or placental origin.

**Table 5.** Common nucleotide point mutations in the HBV preS-S sequence shared between 4 infants born to HBsAg-positive mothers and umbilical cord blood cells which diverged from HBV sequence carried in their mothers.

| Mother-infant pair | Common point mutations |  |
| --- | --- | --- |
|  | Number | Nucleotide substitution at position<sup>a</sup> |
| 9, 10, 11 and 8<sup>b</sup> | 21 | T3242A, G3258A, A3264G, T3270C, C3291A, C3311T, A3328T, C136T, A149G, C151T, C166T, A167C, G191T, A242G, G288A, A290T, C291A, A294G, C295T, C301T, T358C |
| 10, 11 and 8<sup>b</sup> (but not 9) | 4 | C3216T, A3225C, G3235A, A3324T |
| 10 and 8<sup>b</sup> (but not 9 and 11) | 2 | C3246T, C3267T |

<sup>a</sup> Nucleotide substitution comparing to the maternal HBV preS-S sequence spanning between nucleotides 3214 and 359. Nucleotide positions were assigned according to the HBV sequence with GenBank accession number LC057377. For details see the Result section.
<sup>b</sup> Umbilical cord blood mononuclear cells were not available for analysis from Pair 8. However, since the sequence data for the child and mother of Pair 8 were consistent with those for children and mothers from Pairs 9, 10 and 11, they were included here to exemplify overall uniformity of the finding.

## DISCUSSION

The current guidelines for HBV prevention of MTCT recommend HepBvac and HBIG administration to infants from HBsAg-positive mothers within 12 hours after parturition followed by completion of the three-dose HepBvac series [12]. This passive-active prophylaxis has been shown to prevent clinically overt HBV infection in the majority of the newborns [38,39,40], although it failed in 3% to almost 10% of the cases which developed with time serologically evident infection and CHB [9,17,41,42]. In contrast, anti-HBV treatment of HBsAg-positive pregnant women in combination with the perinatal HBV immunoprophylaxis were unable to variable degree halt establishment of OBI in their children [17–20]. To improve outcomes, HepBvac injections beyond the conventional scheme and delivery of additional doses of HBIG were attempted to augment blockage of HBV MTCT [20,40,43]. In the current study, the high-sensitivity HBV detection approach, which was previously successfully used to detect seronegative and seropositive HBV infections and subclinical infections with other hepatitis viruses [5,23–29,35,36], was employed to reassess OBI occurrence among newborns receiving enhanced HBV immune protective measures. The data showed that 84.6% of the children were HBV reactive at low levels during the 24-month follow-up after birth irrespectively of the type of immunoprophylaxis employed. Likewise, antiviral treatment of HBsAg-positive mothers in the last trimester of pregnancy did not contribute to inhibition of HBV MTCT since all infants born to the LdT-treated mothers were HBV DNA reactive at some point during follow-up.

Collectively, the results showed that the preventive measures used had a limited effect on the OBI incidence in newborns from HBsAg-positive mothers. The high rate of OBI incidence uncovered in our study was related to the high-sensitivity HBV detection approach used, which included not only the assay of greater sensitivity, but also testing samples from different compartments and analysis of serial samples collected during the extended follow-up of the children investigated. Applying the same investigation strategy in another study, children born to 48 chronically HBsAg-positive mothers, who were subjected to the standard HBV MTCT preventive protocol and were followed for 12 months after birth, as well as their placentas and umbilical cord blood samples, were examined (Mulrooney-Cousins P.M. *et al.,* unpublished). At 12 months of age, 70.3% of the children were identified as HBV-positive at loads around 1 x 10^2^ vge/mL of serum or plasma. In addition, 61.3% of the placentas examined were HBV DNA reactive, and about 40% of them carried detectable HBV genome replicative intermediates, such as virus RNA and/or covalently closed circular DNA (cccDNA). Regarding umbilical cord blood, HBV DNA was identified at similarly low levels as in children serum or plasma in 64% of the cases tested.

Furthermore, a study employing similarly sensitive virus detection approach was performed in the woodchuck model of hepatitis B in which dams, who resolved experimental acute hepadnaviral hepatitis but carried trace amounts of woodchuck hepatitis virus (WHV) (*i.e.,* < 10^3^ vge/mL of serum) in the presence of otherwise protective anti-virus envelope antibodies, universally transmitted virus to their offspring [25,36]. WHV infection in newborn animals was serum WHV DNA positive but negative for WHV surface antigen (WHsAg; equivalent of HBsAg) and antibodies to WHV antigens, while WHV genome and molecular indicators of its replication were detected in PBMC and organs of the immune system and, in some offspring, in livers [25,36]. This seronegative but molecularly evident WHV infection has been designated as primary occult infection (POI) and extensively examined in the subsequent studies [28,34,36,44-46; reviewed in 3,47,48]. In contrast to POI, spontaneous or therapy-induced resolution of overt WHV infection with hepatitis is invariably followed by lifelong persistence of secondary occult infection (SOI). SOI is serum WHsAg-negative, but positive for WHV DNA, antibodies to WHV core antigen (anti-WHc; equivalent of anti-HBc in HBV infection), while antibodies to WHsAg (anti-WHs; equivalent of anti-HBs) can be transiently detectable during lifelong follow-up [24,47–49]. Notably, WHV in the course of both POI and SOI demonstrates biophysical properties of complete virions and causes classical acute hepatitis and HCC in healthy animals after concentration following ultracentrifugation [5,24,28,36,47]. Because on strong molecular and pathogenic similarities between WHV and HBV, the woodchuck-WHV infection model provides invaluable insights into HBV infection mechanisms and pathogenesis of hepatitis B and HCC, and is successfully used for the preclinical evaluations of novel therapeutics against HBV and the liver diseases caused [48,50,51]. Regarding the relevance of the data from the woodchuck-WHV model to the current study, the immunovirological and molecular HBV infection profiles identified in children indicate that the majority of them acquired OBI compatible with seropositive SOI. However, this HBV DNA-positive, HBsAg-negative and anti-HBc reactive infection became evident only after clearance of maternal HBV-specific antibodies. This suggests that the newborns were exposed to relatively high HBV doses at some point during gestation, likely before initiation of anti-HBV LdT therapy in mothers. Nonetheless, it should be acknowledged that the prophylactic measures employed in our study, although failed to block OBI, suppressed the arise of overt HBV infection for at least two years after birth.

Two of the infants born to DAA-untreated mothers, who carried low HBV loads during pregnancy, did not establish occult infection, as assumed from the repeatedly undetectable HBV DNA by the high-sensitivity approach used. However, both of them became anti-HBc reactive at the age of 2 years. This may suggest that they had seronegative OBI, an equivalent of WHV POI, but carried HBV at levels not detectable by otherwise very sensitive testing or they were *de novo* infected with HBV sometime closely to the end of the observation period. In this regard, the detection of isolated anti-HBc (also called anti-HBc alone) in the absence of HBsAg and in the presence or temporal absence of detectable HBV DNA is recognized as an indicator of protracted low-level HBV infection [52]. Persistence of these antibodies is considered to be indicative of repeated stimulation of the immune system by a trace synthesis of viral nucleocapsid. Experiments in the woodchuck model confirmed that isolated anti-WHc are indicative of SOI that can be accompanied by minimal to moderate hepatitis and could advance to HCC [24,36,48]. Since trans-placentally acquired maternal antibodies wane over the first 6 months of life, anti-HBc detected for the first time in the 2-year-old infants have to be produced in response to either *de novo* HBV infection or reactivation of ongoing seronegative POI. However, there also is a third possibility, the data from the woodchuck model informs that animals with POI remain susceptible to reinfection with high doses (>10^3^ virions) of WHV and in response produce anti-WHc, as well as reactivate virus-specific T cell response [28,44]. Thus, at this stage, it remains unclear why exactly these two children became anti-HBc reactive so late after birth.

Despite a very low-level HBV infection in the children and in some of their mothers, the generation of HBV preS-S gene amplicons and their subsequent sequencing turned out to be feasible. The HBV preS-S fragment was selected for analysis because of its predisposition to mutations and forthright applicability for HBV genotyping. As the data showed, there was unexpected homology between the preS-S sequence found in the children and mononuclear cells of umbilical cord blood that diverged from the corresponding maternal sequences occurring in peripheral mononuclear cells and/or plasma. This was a compelling finding since it was encountered in 4 of the 5 mother-child pairs examined. This prompted search for the possible origin of such HBV genomic discrepancy based upon the data reported and our findings. In this regard, it has been shown that maternal blood mononuclear cells infected with HBV can cross the placental barrier, and facilitate virus intrauterine transmission and fetus infection [53–57]. This agrees with the two-way cell trafficking between mother and fetus across the barrier [58]. It also is established that PBMC infected with HBV, as well as with WHV, can support active virus replication and become a reservoir of the infectious and liver pathogenic virus [3,21,59,60]. Due to the hepadnaviral replication strategy, which engages the low fidelity transcription of HBV pregenomic RNA to virus DNA, HBV is highly prone to mutations resulting in multiple HBV subtypes, variants and in compartment sequence diversity [61,62]. Thus, it can be expected that HBV replicating in different milieus in a mother, fetus and placenta is prone to creation of distinct HBV variants, including those carried within circulating mononuclear immune cells. Since the traffic of cells from the fetus to mother via the cord blood compartment is known [58,63], the uniformity of HBV variants in newborns and umbilical cord blood cells, as it is exemplified by the detected HBV variants in our study, is highly feasible. There is so far a singular study comparing HBV quasispecies diversity between infants with OBI and their viremic mothers, although virus genomes derived only from sera were investigated [64].

Nonetheless, the analysis of the HBV whole sequences showed overall increased complexity of point mutations in infants compared to their mothers. It is rather unlikely that the HBV sequence diversified in the period between birth and the sample collection at 7 and 12 months of age and, therefore, suggests that this process occurred during the gestation period. Another study examined HBV genetic diversity in maternal and cord blood plasma and in the placenta samples [65]. The data implied the lack of unique mutations in these three types of samples from the same cases and suggested that mutations were unlikely to occur during intrauterine exposure to HBV. In opposite, our study focused on examination of the 360-bp preS-S fragment was able to identify uniform HBV mutation when cells from umbilical cord blood and whole blood samples from children with OBI were investigated.

This indicates that circulating mononuclear cells should be analyzed when searching for origins of HBV variants found in children born to HBV-infected mothers. The variant uniformity uncovered in our study can be interpreted in support of two notions: (1) circulating mononuclear cells transmit infectious HBV during the fetal development and (2) either placenta or fetus or both are the sites where mononuclear immune cells may acquire mutated HBV. More detailed investigations of the maternal PBMC may identify the variant matching that found in umbilical cord blood cells and children, but this will require a separate study. Regarding that, this also needs to be considered that HBV variants may reside in a particular immune cell subset and may not be detectable in total PBMC, as it was reported for occult HCV infection [66,67].

The possible placental origin of the HBV preS-S variant detected in our study cannot be excluded, although we did not have in our hands appropriate materials to confirm or discount this possibility. In this context, HBV infection of trophoblast cells and possibly other cells in placenta has been reported [52,68,69]. Indirect but relevant evidence also comes from infections with other viruses, including transmission of human immunodeficiency virus type 1 between T cells and placental trophoblast cells [70,71]. Since HBV also is a lymphotropic virus [3,72], a similar route of viral spreading may operate within placentas of pregnant women infected with HBV.

By applying the high-sensitivity HBV detection approach, our findings document the much higher incidence of subclinical HBV infection in infants born to HBsAg-positive mothers than previously reported. The virus persisted for up to the 2-year follow-up after birth, although all infants received enhanced HBV immunoprophylaxis. The treatment of mothers with HBV-specific DAA in the third trimester of pregnancy did not cease development of OBI. Taken together, our findings confirmed that the current approach to terminate HBV MTCT does not stop transfer of HBV traces capable of initiating clinically silent infection in newborns. Infection of fetus early in pregnancy, prior to initiation of maternal anti-HBV therapy in the third semester, might be one of the reasons why OBI was established.

However, based on yet limited clinical observations and considering data from the woodchuck-WHV model of OBI, it appears that transmission of HBV traces throughout the placental barrier, fetus infection, and subsequent establishment of occult infection in children could be an inevitable consequence of maternal exposure to HBV. The long-term pathogenic outcomes of such acquired infection might not be clinically obvious or numerous and recognition of their full scope will require much more investigation using augmented virus detection techniques. However, the documented ability of HBV, as well as traces of WHV [28], to integrate into the host’s genome almost immediately after infection and persistence of potentially oncogenic virus-host genome integrations may have serious life-treating consequences [28,51,73,74]. Since there is not currently therapy to completely eliminate HBV from the infected host, our study emphasizes a tremendous importance of global HBV vaccination, in particular vaccination of infants and young adults, to avert virus vertical spread and protect future generations.

## Data Availability

All data produced in the present work are contained in the manuscript.

## Acknowledgements

The authors thanks Norma D. Churchill from the Molecular Virology and Hepatology Research Group, Faculty of Medicine, Memorial University, St. John’s, NL, Canada for reviewing manuscript draft.

## List of Abbreviations

ACD: acid-citrate-dextrose
ALP: alkaline phosphatase
ALT: alanine aminotransferase
anti-HBc: antibodies to HBV core antigen
anti-HBe: antibodies to HBV e antigen
anti-HBs: antibodies to HBV surface antigen
anti-WHc: antibodies to WHV core antigen
anti-WHs: antibodies to WHV surface antigen
cccDNA: covalently closed circular DNA
CHB: chronic hepatitis B
COI: cut-off index
DAA: directly acting antivirals
HBeAg: HBV e antigen
HBIG: hepatitis B immunoglobulin
HBsAg: HBV surface antigen
HBV: hepatitis B virus
HBV C: HBV core (nucleocapsid) gene
HBV preS-S: HBV preS-S fragment of the HBV S gene
HBV S: HBV surface (envelope) gene
HBV X: HBV X gene
HCC: hepatocellular carcinoma
HepBvac: HBV vaccine
IFTM group: infant-from-treated-mother group
IFUTM group: infant-from-untreated-mother group
MHVL group: maternal high virus load group
MLVL group: maternal low virus load group
MPBC or MC: maternal peripheral blood mononuclear cells or maternal cells
MPBP or MP: maternal peripheral blood plasma or maternal plasma
MTCT: mother-to-child transmission
NAH: nucleic acid hybridization
OBI: occult HBV infection
PCR: polymerase chain reaction
POI: primary (seronegative) occult infection
SOI: secondary (seropositive) occult WHV infection
UCBP: umbilical cord blood plasma
UCBC or UC: umbilical cord blood mononuclear cells or umbilical cells
vge: virus genome equivalents
WBC: whole blood clot
WBC02 or B02: whole blood clot from newborn at 2 days of life
WBC2 or B2: whole blood clot from 2 months old infant
WBC7 or B7: whole blood clot from 7 months old infant
WBC12 or B12: whole blood clot from 12 months old infant
WBC24 or B24: whole blood clot from 24 months old infant
WHV: woodchuck hepatitis virus

## References

1. World Health Organization Hepatitis B Fact Sheet. Updated April 2024. World Health Organization (2024). Geneva, Switzerland (accessed on 09 November 2024).

2. Raimondo G, Locarnini S, Pollicino T, Levrero M, Zoulim F, Lok AS; Taormina Workshop on Occult HBV Infection Faculty Members. Update of the statements on biology and clinical impact of occult hepatitis B virus infection. J Hepatol. 2019;71:397–408. doi: 10.1016/j.jhep.2019.03.034.

3. Coffin CS, Mulrooney-Cousins PM, Michalak TI. Hepadnaviral lymphotropism and its relevance to HBV persistence and pathogenesis. Front Microbiol. 2021;12:695384. doi: 10.3389/fmicb.2021.695384.

4. Blum HE, Liang TJ, Galun E, Wands JR. Persistence of hepatitis B viral DNA after serological recovery from hepatitis B virus infection. Hepatology. 1991;14:56–63. doi: 10.1002/hep.1840140110.

5. Michalak TI, Pasquinelli C, Guilhot S, Chisari FV. Hepatitis B virus persistence after recovery from acute viral hepatitis. J Clin Invest. 1994;93:230–239. doi: 10.1172/JCI116950.

6. Mulrooney-Cousins PM, Michalak TI. Asymptomatic hepadnaviral persistence and Its consequences in the woodchuck model of occult hepatitis B virus infection. J Clin Transl Hepatol. 2015;3:211–219. doi: 10.14218/JCTH.2015.00020.

7. Saitta C, Pollicino T, Raimondo G. Occult hepatitis B virus infection: An update. Viruses. 2022;14:1504. doi: 10.3390/v14071504.

8. Candotti D, Assennato SM, Laperche S, Allain JP, Levicnik-Stezinar S. Multiple HBV transfusion transmissions from undetected occult infections: revising the minimal infectious dose. Gut. 2019;68:313–321. doi: 10.1136/gutjnl-2018-316490.

9. Eilard A, Andersson M, Ringlander J, Wejstål R, Norkrans G, Lindh M. Vertically acquired occult hepatitis B virus infection may become overt after several years. J Infect. 2019;78:226–231. doi: 10.1016/j.jinf.2019.01.002.

10. Mak LY, Wong DK, Pollicino T, Raimondo G, Hollinger FB, Yuen MF. Occult hepatitis B infection and hepatocellular carcinoma: Epidemiology, virology, hepatocarcinogenesis and clinical significance. J Hepatol. 2020;73:952–964. doi: 10.1016/j.jhep.2020.05.042.

11. Shi Y, Zheng M. Hepatitis B virus persistence and reactivation. BMJ. 2020;370:m2200. Doi: 10.1136/bmj.m2200.

12. Guidelines for the prevention, diagnosis, care and treatment for people with chronic hepatitis B infection. World Health Organization (2024). Geneva, Switzerland.

13. Liu Z, Chen Z, Cui F, Ding Y, Gao Y, Han G, Jia J, Li J, Li Z, Liu Y, Mao Q, Wang A, Wang W, Wei L, Xia J, Xie Q, Yang X, Yin X, Zhang H, Zhang L, Zhang W, Zhuang H, Dou X, Hou J. Management algorithm for prevention of mother-to-child transmission of hepatitis B virus (2022). J Clin Transl Hepatol. 2022;10:1004–1010. doi: 10.14218/JCTH.2022.00047.

14. Shahmoradi S, Yahyapour Y, Mahmoodi M, Alavian SM, Fazeli Z, Jazayeri SM. High prevalence of occult hepatitis B virus infection in children born to HBsAg-positive mothers despite prophylaxis with hepatitis B vaccination and HBIG. J Hepatol. 2012;57:515–521. doi: 10.1016/j.jhep.2012.04.021.

15. Su H, Zhang Y, Xu D, Wang B, Zhang L, Li D, Xiao D, Li F, Zhang J, Yan Y. Occult hepatitis B virus infection in anti-HBs-positive infants born to HBsAg-positive mothers in China. PloS One. 2013;8:e70768. doi: 10.1371/journal.pone.0070768.

16. Foaud H, Maklad S, Mahmoud F, El-Karaksy H. Occult hepatitis B virus infection in children born to HBsAg-positive mothers after neonatal passive-active immunoprophylaxis. Infection. Infection. 2015;43:307–314. doi: 10.1007/s15010-015-0733-6.

17. Pande C, Sarin SK, Patra S, Kumar A, Mishra S, Srivastava S, Bhutia K, Gupta E, Mukhopadhyay CK, Dutta AK, Trivedi SS. Hepatitis B vaccination with or without hepatitis B immunoglobulin at birth to babies born of HBsAg-positive mothers prevents overt HBV transmission but may not prevent occult HBV infection in babies: a randomized controlled trial. J Viral Hepat. 2013;20:801–810. doi: 10.1111/jvh.12102.

18. Lu Y, Liu YL, Nie JJ, Liang XF, Yan L, Wang FZ, Zhai XJ, Liu JX, Zhu FC, Chang ZJ, Li J. Occult HBV infection in immunized neonates born to HBsAg-positive mothers: A prospective and follow-up study. PloS One. 2016;11:e0166317. Doi: 10.1371/journal.pone.0166317.

19. Zhou S, Li T, Allain JP, Zhou B, Zhang Y, Zhong M, Fu Y, Li C. Low occurrence of HBsAg but high frequency of transient occult HBV infection in vaccinated and HBIG-administered infants born to HBsAg positive mothers. J Med Virol. 2017;89:2130–2137. doi: 10.1002/jmv.24861.

20. Li Y, Li L, Song Y, Liu M, Zhai X, Duan Z, Ding F, Zhu L, Jiang J, Zou H, Wang J, Li J. Booster vaccination in infancy reduces the incidence of occult HBV infection in maternal HBsAg-positive children. J Clin Transl Hepatol. 2023;11:661–669. doi: 10.14218/JCTH.2022.00213.

21. Coffin CS, Mulrooney-Cousins PM, Peters MG, van Marle G, Roberts JP, Michalak TI, Terrault NA. Molecular characterization of intrahepatic and extrahepatic hepatitis B virus (HBV) reservoirs in patients on suppressive antiviral therapy. J Viral Hepat. 2011;18:415–423. doi: 10.1111/j.1365-2893.2010.01321.x.

22. Coffin CS, Mulrooney-Cousins PM, van Marle G, Roberts JP, Michalak TI, Terrault NA. Hepatitis B virus quasispecies in hepatic and extrahepatic viral reservoirs in liver transplant recipients on prophylactic therapy. Liver Transpl. 2011;17:955–962. doi: 10.1002/lt.22312.

23. Skardasi G, Chen AY, Michalak TI. Authentic patient-derived hepatitis C virus infects and productively replicates in primary CD4^+^ and CD8^+^ T lymphocytes *in vitro*. J Virol. 2018;92:e01790–17. doi: 10.1128/JVI.01790-17.

24. Michalak TI, Pardoe IU, Coffin CS, Churchill ND, Freake DS, Smith P, Trelegan CL. Occult lifelong persistence of infectious hepadnavirus and residual liver inflammation in woodchucks convalescent from acute viral hepatitis. Hepatology. 1999;29:928–938. doi: 10.1002/hep.510290329.

25. Coffin CS, Michalak TI. Persistence of infectious hepadnavirus in the offspring of woodchuck mothers recovered from viral hepatitis. J Clin Invest. 1999;104:203–212. doi: 10.1172/JCI5048.

26. Pham TN, MacParland SA, Mulrooney PM, Cooksley H, Naoumov NV, Michalak TI. Hepatitis C virus persistence after spontaneous or treatment-induced resolution of hepatitis C. J Virol. 2004;78:5867–5874. doi: 10.1128/JVI.78.11.5867-5874.2004.

27. Pham TN, King D, MacParland SA, McGrath JS, Reddy SB, Bursey FR, Michalak TI. Hepatitis C virus replicates in the same immune cell subsets in chronic hepatitis C and occult infection. Gastroenterology. 2008;134:812–822. doi: 10.1053/j.gastro.2007.12.011.

28. Mulrooney-Cousins PM, Chauhan R, Churchill ND, Michalak TI. Primary seronegative but molecularly evident hepadnaviral infection engages liver and induces hepatocarcinoma in the woodchuck model of hepatitis B. PLoS Pathog. 2014;10(8):e1004332. doi: 10.1371/journal.ppat.1004332.

29. Chen AY, Hoare M, Shankar AN, Allison M, Alexander GJ, Michalak TI. Persistence of hepatitis C virus traces after spontaneous resolution of hepatitis C. PloS One. 2015;10:e0140312. doi: 10.1371/journal.pone.0140312.

30. Joshi D, James A, Quaglia A, Westbrook RH, Heneghan MA. Liver disease in pregnancy. Lancet. 2010;375:594–605. doi: 10.1016/S0140-6736(09)61495-1.

31. Ruhl CE, Everhart JE. Upper limits of normal for alanine aminotransferase activity in the United States population. Hepatology. 2012;55:447–454. doi: 10.1002/hep.24725.

32. Pardoe IU, Michalak TI. Detection of hepatitis B and woodchuck hepatitis viral DNA in plasma and mononuclear cells from heparinized blood by the polymerase chain reaction. J Virol Methods. 1995;51(2-3):277–288. doi: 10.1016/0166-0934(94)00116-x.

33. Demeke T, Jenkins GR. Influence of DNA extraction methods, PCR inhibitors and quantification methods on real-time PCR assay of biotechnology-derived traits. Anal Bioanal Chem. 2010;396:1977–1990. doi: 10.1007/s00216-009-3150-9.

34. Corkum CP, Ings DP, Burgess C, Karwowska S, Kroll W, Michalak TI. Immune cell subsets and their gene expression profiles from human PBMC isolated by Vacutainer Cell Preparation Tube (CPT™) and standard density gradient. BMC Immunol. 2015;16:48. doi: 10.1186/s12865-015-0113-0.

35. Michalak TI. Occult persistence and lymphotropism of hepadnaviral infection: insights from the woodchuck viral hepatitis model. Immunol Rev. 2000;174:98–111. doi: 10.1034/j.1600-0528.2002.017406.x.

36. Michalak TI, Mulrooney PM, Coffin CS. Low doses of hepadnavirus induce infection of the lymphatic system that does not engage the liver. J Virol. 2004;78:1730–1738. doi: 10.1128/jvi.78.4.1730-1738.2004.

37. Pham TN, Michalak TI. Occult persistence and lymphotropism of hepatitis C virus infection. World J Gastroenterol. 2008;14:2789–2793. doi: 10.3748/wjg.14.2789.

38. Lee C, Gong Y, Brok J, Boxall EH, Gluud C. Effect of hepatitis B immunization in newborn infants of mothers positive for hepatitis B surface antigen: Systematic review and meta-analysis. BMJ. 2006;332:328–336. doi: 10.1136/bmj.38719.435833.7C.

39. Machaira M, Papaevangelou V, Vouloumanou EK, Tansarli GS, Falagas ME. Hepatitis B vaccine alone or with hepatitis B immunoglobulin in neonates of HBsAg+/HBeAg-mothers: A systematic review and meta-analysis. J Antimicrob Chemother. 2015;70:396–404. doi: 10.1093/jac/dku404.

40. Sintusek P, Wanlapakorn N, Poovorawan Y. Strategies to prevent mother-to-child transmission of hepatitis B virus. J Clin Transl Hepatol. 2023;11:967–974. doi: 10.14218/JCTH.2022.00332.

41. Chen HL, Lin LH, Hu FC, Lee JT, Lin WT, Yang YJ, Huang FC, Wu SF, Chen SC, Wen WH, Chu CH, Ni YH, Hsu HY, Tsai PL, Chiang CL, Shyu MK, Lee PI, Chang FY, Chang MH. Effects of maternal screening and universal immunization to prevent mother-to-infant transmission of HBV. Gastroenterology. 2012;142:773–781.e2. doi: 10.1053/j.gastro.2011.12.035.

42. Lin X, Guo Y, Zhou A, Zhang Y, Cao J, Yang M, Xiao F, Zhang B, Du Y. Immunoprophylaxis failure against vertical transmission of hepatitis B virus in the Chinese population: a hospital-based study and a meta-analysis. Pediatr Infect Dis J. 2014;33:897–903. doi: 10.1097/INF.0000000000000315.

43. Poovorawan Y, Sanpavat S, Chumdermpadetsuk S, Safary A. Long-term hepatitis B vaccine in infants born to hepatitis B e antigen positive mothers. Arch Dis Child Fetal Neonatal Ed. 1997;77:F47–51. doi: 10.1136/fn.77.1.f47.

44. Gujar SA, Michalak TI. Primary occult hepadnavirus infection induces virus-specific T-cell and aberrant cytokine responses in the absence of antiviral antibody reactivity in the Woodchuck model of hepatitis B virus infection. J Virol. 2009;83:3861–3876. doi: 10.1128/JVI.02521-08.

45. Gujar SA, Mulrooney-Cousins PM, Michalak TI. Repeated exposure to trace amounts of woodchuck hepadnavirus induces molecularly evident infection and virus-specific T cell response in the absence of serological infection markers and hepatitis. J Virol. 2013;87:1035–1048. doi: 10.1128/JVI.01363-12.

46. Williams JB, Hüppner A, Mulrooney-Cousins PM, Michalak TI. Differential Expression of Woodchuck Toll-Like Receptors 1-10 in Distinct Forms of Infection and Stages of Hepatitis in Experimental Hepatitis B Virus Infection. Front Microbiol. 2018;9:3007. doi: 10.3389/fmicb.2018.03007.

47. Michalak TI. Diverse virus and host-dependent mechanisms influence the systemic and intrahepatic immune responses in the woodchuck model of hepatitis B. Front Immunol. 2020;11:853. doi: 10.3389/fimmu.2020.00853.

48. Michalak TI. Immunological aspects of naturally occurring model of HBV Infection, hepatitis B and HBV-associated hepatocellular carcinoma in the American woodchuck *Marmota monax*. J Immunol. Oct 2025 (in press).

49. Coffin CS, Pham TN, Mulrooney PM, Churchill ND, Michalak TI. Persistence of isolated antibodies to woodchuck hepatitis virus core antigen is indicative of occult infection. Hepatology. 2004;40:1053–1061. doi: 10.1002/hep.20419

50. Suresh M, Menne S. Recent drug development in the woodchuck model of chronic hepatitis B. Viruses. 2022;14:1711. doi: 10.3390/v14081711.

51. Chauhan R, Churchill ND, Mulrooney-Cousins PM, Michalak TI. Initial sites of hepadnavirus integration into host genome in human hepatocytes and in the woodchuck model of hepatitis B-associated hepatocellular carcinoma. Oncogenesis. 2017;6:e317. doi: 10.1038/oncsis.2017.2.

52. Wang Q, Klenerman P, Semmo N. Significance of anti-HBc alone serological status in clinical practice. Lancet Gastroenterol Hepatol. 2017;2:123–134. doi: 10.1016/S2468-1253(16)30076-0.

53. Bai H, Zhang L, Ma L, Dou XG, Feng GH, Zhao GZ. Relationship of hepatitis B virus infection of placental barrier and hepatitis B virus intra-uterine transmission mechanism. World J Gastroenterol. 2007;13:3625–3630. doi: 10.3748/wjg.v13.i26.3625.

54. Shao Q, Zhao X, Yao Li MD. Role of peripheral blood mononuclear cell transportation from mother to baby in HBV intrauterine infection. Arch Gynecol Obstet. 2013;288:1257–1261. doi: 10.1007/s00404-013-2893-x.

55. Xu YY, Liu HH, Zhong YW, Liu C, Wang Y, Jia LL, Qiao F, Li XX, Zhang CF, Li SL, Li P, Song HB, Li Q. Peripheral blood mononuclear cell traffic plays a crucial role in mother-to-infant transmission of hepatitis B virus. Int J Biol Sci. 2015;1:266–273. doi: 10.7150/ijbs.10813.

56. Shi X, Wang X, Xu X, Feng Y, Li S, Feng S, Wang B, Wang S. Impact of HBV replication in peripheral blood mononuclear cell on HBV intrauterine transmission. Front Med. 2017;11:548–553. doi: 10.1007/s11684-017-0597-5.

57. Wang DD, Yi LZ, Wu LN, Yang ZQ, Hao HY, Shi XH, Wang B, Feng SY, Feng YL, Wang SP. Relationship between Maternal PBMC HBV cccDNA and HBV Serological Markers and its Effect on HBV Intrauterine Transmission. Biomed Environ Sci. 2019;32:315–323. doi: 10.3967/bes2019.043.

58. Lo YM, Lo ES, Watson N, Noakes L, Sargent IL, Thilaganathan B, Wainscoat JS. Two-way cell traffic between mother and fetus: biologic and clinical implications. Blood. 1996;88:4390–4395. PMID: 8943877.

59. Lu L, Zhang HY, Yueng YH, Cheung KF, Luk JM, Wang FS, Lau GK. Intracellular levels of hepatitis B virus DNA and pregenomic RNA in peripheral blood mononuclear cells of chronically infected patients. J Viral Hepat. 2009;16:104–112. doi: 10.1111/j.1365-2893.2008.01054.x.

60. Gao S, Duan ZP, Chen Y, van der Meer F, Lee SS, Osiowy C, van Marle G, Coffin CS. Compartmental HBV evolution and replication in liver and extrahepatic sites after nucleos/tide analogue therapy in chronic hepatitis B carriers. J Clin Virol. 2017;94:8–14. doi: 10.1016/j.jcv.2017.06.009.

61. Glebe D, Goldmann N, Lauber C, Seitz S. HBV evolution and genetic variability: Impact on prevention, treatment and development of antivirals. Antiviral Res. 2021;186:104973. doi: 10.1016/j.antiviral.2020.104973.

62. Xie C, Lu D. Evolution and diversity of the hepatitis B virus genome: Clinical implications. Virology. 2024;598:110197. doi: 10.1016/j.virol.2024.110197.

63. Lo YM, Lau TK, Chan LY, Leung TN, Chang AM. Quantitative analysis of the bidirectional fetomaternal transfer of nucleated cells and plasma DNA. Clin Chem. 2000;46:1301–1309.

64. Li Y, Song Y, Xiao Y, Wang T, Li L, Liu M, Li J, Wang J. The Characteristic of HBV quasispecies is related to occult HBV infection of infants born to highly viremic mothers. Viruses. 2024;16:1104. doi: 10.3390/v16071104.

65. Sirilert S, Khamrin P, Kumthip K, Malasao R, Maneekarn N, Tongsong T. Possible Association between genetic diversity of hepatitis B virus and its effect on the detection rate of hepatitis B virus DNA in the placenta and fetus. Viruses. 2023;15:1729. doi: 10.3390/v15081729.

66. Pham TN, King D, MacParland SA, McGrath JS, Reddy SB, Bursey FR, Michalak TI. Hepatitis C virus replicates in the same immune cell subsets in chronic hepatitis C and occult infection. Gastroenterology. 2008;134:812–822. doi: 10.1053/j.gastro.2007.12.01.

67. Michalak TI, Pham TNQ, Mulrooney-Cousins PM. Molecular diagnosis of occult HCV and HBV infections. Future Virol. 2007;2:241–246. doi: 10.2217/17460794.2.5.451.

68. Xu DZ, Yan YP, Choi BC, Xu JQ, Men K, Zhang JX, Liu ZH, Wang FS. Risk factors and mechanism of transplacental transmission of hepatitis B virus: a case-control study. J Med Virol. 2002;67:20–26. doi: 10.1002/jmv.2187.

69. Zhao X, Bai X, Xi Y. Intrauterine infection and mother-to-child transmission of hepatitis B virus: Route and molecular mechanism. Infect Drug Resist. 2022;15:1743–1751. doi: 10.2147/IDR.S359113.

70. Arias RA, Muñoz LD, Muñoz-Fernández MA. Transmission of HIV-1 infection between trophoblast placental cells and T-cells take place via an LFA-1-mediated cell to cell contact. Virology. 2003;307:266–277. doi: 10.1016/s0042-6822(02)00040-5

71. Lagaye S, Derrien M, Menu E, Coïto C, Tresoldi E, Mauclère P, Scarlatti G, Chaouat G, Barré-Sinoussi F, Bomsel M; European Network for the Study of In Utero Transmission of HIV-1. Cell-to-cell contact results in a selective translocation of maternal human immunodeficiency virus type 1 quasispecies across a trophoblastic barrier by both transcytosis and infection. J Virol. 2001;75:4780-4791. doi: 10.1128/JVI.75.10.4780-4791.2001.

72. Svicher V, Salpini R, D’Anna S, Piermatteo L, Iannetta M, Malagnino V, Sarmati L. New insights into hepatitis B virus lymphotropism: Implications for HBV-related lymphomagenesis. Front Oncol. 2023;13:1143258. doi: 10.3389/fonc.2023.1143258.

73. Tu T, Budzinska MA, Shackel NA, Urban S. HBV DNA integration: molecular mechanisms and clinical implications. Viruses. 2017;9:75. doi:10.3390/v9040075

74. Michalak TI. The initial hepatitis B virus-hepatocyte genomic integrations and their role in hepatocellular oncogenesis. Int J Mol Sci. 2023;24:14849. doi: 10.3390/ijms241914849.

