## supplementary Table 1 for "Hepatitis B Prophylaxis Rarely Prevents Subclinical Molecularly Evident HBV Infection in Children Born to HBsAg-Positive Mothers"

**Supplementary Table 1.** Oligonucleotide primers used for HBV DNA amplification by PCR

| Gene | Reaction | Primer | Location (nt)* | Sequence | Sense/Antisense |
| --- | --- | --- | --- | --- | --- |
| S | dPCR | SDP | 2816-2838 | 5'-TTG TGG GTC ATA TTC TTG G-3' | sense |
|  |  | SDM | 450-472 | 5'-CGG GCA ACA TAC CTT GAT AGT CC-3' | antisense |
|  | nPCR | SNP | 3057-3078 | 5'-ATC CTC AGG CCA TGC AGT GG-3' | sense |
|  |  | SNM | 365-387 | 5'-GCA GAC ACA TCC AGC GAT AAC C-3' | antisense |
| X | dPCR | XDP | 1248-1288 | 5'-CCA TAC TGC GGA ACT CCT AGC-3' | sense |
|  |  | XDM | 1781-1804 | 5'-ACA GAC CAA TTT ATG CCT ACA GCC-3' | antisense |
|  | nPCR | XNP | 1312-1331 | 5'-CTG GAG CAA ACA TTA TCG GG-3' | sense |
|  |  | XNM | 1750-1772 | 5'-CAA AGA CCT TTA ACC TGA TCT CC-3' | antisense |
| C | dPCR | CDP | 1849-1871 | 5'-TGT TCA TGT CCC ACT GTT CAA GC-3' | sense |
|  |  | CDM | 2302-2321 | 5'-AAG ATA GGG GCA TTT GGT GG-3' | antisense |
|  | nPCR | CNP | 1895-1915 | 5'-TTT GGG GCA TGG ACA TTG ACC-3' | sense |
|  |  | CNM | 2276-2298 | 5'-ATA AGC TGG AGG AGT GCG AAT CC-3' | antisense |

*Abbreviations:* nt, nucleotide number; dPCR, direct PCR; nPCR, nested PCR.

* Nucleotide positions according to the HBV DNA sequence under GenBank accession number LC057377.
