## Supplementary Table 3 for "Hepatitis B Prophylaxis Rarely Prevents Subclinical Molecularly Evident HBV Infection in Children Born to HBsAg-Positive Mothers"

**Supplementary Table 3.** Immunovirological and molecular markers of HBV infection in HBIG-treated, HBV-vaccinated infants born to HBsAg-positive mothers at 2 days and 2, 7, 12 and 24 months of life

| Infant  number | 2 days | | | | | |  | 2 months | | | | | |  | 7 months | | | | | |
| --- | --- | --- | --- | --- | --- | --- | --- | --- | --- | --- | --- | --- | --- | --- | --- | --- | --- | --- | --- | --- |
|  | HBsAg  (IU/mL)^a^ | Anti-HBs  (mIU/mL)^a^ | HBeAg  (COI) ^a^ | Anti-HBe  (COI)^a^ | Anti-HBc  (COI)^a^ | HBV DNA (vge/mL)^b^ |  | HBsAg  (IU/mL)^a^ | Anti-HBs  (mIU/mL)^a^ | HBeAg  (COI)^a^ | Anti-HBe  (COI)^a^ | Anti-HBc  (COI)^a^ | HBV DNA (vge/mL)^b^ |  | HBsAg  (IU/mL)^a^ | Anti-HBs  (mU/mL)^a^ | HBeAg  (COI)^a^ | Anti-HBe  (COI)^a^ | Anti-HBc  (COI)^a^ | HBV DNA  (vge/mL)^b^ |
| Infant 1/M | neg (<0.05) | neg (<10.0) | neg (0.06) | pos (0.007) | pos (0.012) | nt |  | neg (<0.05) | pos (272.1) | neg (0.14) | pos (0.025) | pos (0.008) | nt |  | neg (<0.05) | pos (996.0) | neg (0.12) | neg (1.3) | pos (0.187) | neg |
| Infant 2/M | neg (<0.05) | neg (<10.0) | neg (0.11) | pos (0.003) | pos (0.008) | nt |  | neg (<0.05) | pos (159.6) | neg (0.11) | pos (0.003) | pos (0.010) | nt |  | neg (<0.05) | neg (6.3) | neg (0.12) | pos (0.464) | pos (0.007) | neg |
| Infant 3/F | neg (<0.05) | neg (<10.0) | neg (0.11) | pos (0.019) | pos (0.006) | nt |  | neg (<0.05) | pos (316.2) | neg (0.11) | pos (0.061) | pos (0.007) | nt |  | neg (<0.05) | pos (616.6) | neg (0.13) | pos (0.488) | pos (0.013) | 1.2 x 10^2^ |
| Infant 4/F | neg (<0.05) | neg (<10.0) | neg (0.10) | pos (0.007) | pos (0.006) | nt |  | neg (<0.05) | pos (358.8) | neg (0.11) | pos (0.044) | pos (0.007) | neg |  | neg (<0.05) | pos (>1000) | neg (0.12) | pos (0.88) | pos (0.005) | 8.1 x 10^1^ |
| Infant 5/M | neg (<0.05) | neg (<10.0) | pos (236.9) | neg (2.03) | pos (0.008) | nt |  | neg (<0.05) | pos (302.8) | pos (5.72) | neg (1.34) | pos (0.009) | neg |  | neg (<0.05) | pos (620.5) | neg (0.08) | neg (1.38) | pos (0.007) | 1.1 x10^2^ |
| Infant 6/M | neg (<0.05) | neg (<10.0) | pos (125.2) | neg (1.42) | pos (0.010) | nt |  | neg (<0.05) | pos (364.2) | pos (3.55) | neg (1.44) | pos (0.020) | nt |  | neg (<0.05) | pos (>1000) | neg (0.33) | neg (1.52) | pos (0.048) | nt |
| Infant 7/M | neg (<0.05) | neg (<10.0) | neg (0.07) | pos (0.281) | pos (0.013) | nt |  | neg (<0.05) | pos (299.5) | neg (0.08) | neg (1.08) | pos (0.017) | nt |  | neg (<0.05) | pos (667.4) | neg (0.24) | neg (1.59) | pos (0.023) | nt |
| Infant 8/F | neg (<0.05) | pos (20.3) | pos (164.0) | neg (1.9) | pos (0.008) | nt |  | neg (<0.05) | pos (477.8) | neg (0.21) | neg (1.41) | pos (0.010) | 1.1 x 10^2^ |  | neg (<0.05) | pos (392.2) | neg (0.12) | neg (1.62) | pos (0.025) | 1.8 x 10^2^ |
| Infant 9/F | neg (<0.05) | neg (<10.0) | pos (12.3) | neg (1.3) | pos (0.006) | nt |  | neg (<0.05) | pos (454.8) | neg (0.11) | neg (1.26) | pos (0.007) | 1.1 x 10^2^ |  | neg (<0.05) | pos (495.7) | neg (0.27) | neg (1.74) | neg (8.75) | 1.5 x 10^2^ |
| Infant 10/F | neg (<0.05) | neg (<10.0) | pos (146.7) | neg (1.67) | pos (0.007) | nt |  | neg (<0.05) | pos (324.0) | neg (0.14) | neg (1.44) | pos (0.007) | 1.2 x 10^2^ |  | neg (<0.05) | pos (18.81) | neg (0.27) | neg (1.77) | neg (7.81) | 9.7 x 10^2^ |
| Infant 11/F | neg (<0.05) | neg (<10.0) | neg (0.10) | pos (0.003) | pos (0.008) | nt |  | neg (<0.05) | pos (280.7) | neg (0.13) | neg (1.62) | pos (0.016) | 2.1 x 10^2^ |  | neg (<0.05) | pos (178.0) | neg (0.11) | neg (1.81) | pos (0.712) | nt |
| Infant 12/M | neg (<0.05) | pos (14.3) | pos (10.6) | neg (1.73) | pos (0.006) | nt |  | neg (<0.05) | pos (322.1) | neg (0.12) | neg (1.62) | pos (0.007) | 2.0 x 10^2^ |  | neg (<0.05) | pos (16.3) | neg (0.1) | neg (1.76) | pos (0.372) | 3.6 x 10^2^ |
| Infant 13/F | neg (<0.05) | neg (<10.0) | pos (76.9) | neg (1.197) | pos (0.010) | nt |  | neg (<0.05) | pos (544.0) | neg (0.72) | neg (1.30) | pos (0.116) | nt |  | neg (<0.05) | pos (>1000) | neg (0.08) | neg (1.03) | pos (0.027) | nt |
| Positive/  total tested | 0/13 | 2/13 | 7/13 | 6/13 | 13/13 | nt |  | 0/13 | 13/13 | 2/13 | 4/13 | 13/13 | 5/7 |  | 0/13 | 13/13 | 0/13 | 3/13 | 11/13 | 7/9 |

| Infant  number | 12 months | | | | | |  | 24 moths | | | | | |
| --- | --- | --- | --- | --- | --- | --- | --- | --- | --- | --- | --- | --- | --- |
|  | HBsAg  (IU/mL)^a^ | Anti-HBs  (mIU/mL)^a^ | HBeAg  (COI)^a^ | Anti-HBe  (COI)^a^ | Anti-HBc  (COI)^a^ | HBV DNA (vge/mL)^b^ |  | HBsAg  (IU/mL)^a^ | Anti-HBs  (mIU/mL)^a^ | HBeAg  (COI)^a^ | Anti-HBe  (COI)^a^ | Anti-HBc  (COI)^a^ | HBV DNA (vge/mL)^b^ |
| Infant 1/M | neg (<0.05) | pos (193.0) | neg (0.30) | neg (1.55) | pos (0.48) | neg |  | neg (<0.05) | pos (>1000) | neg (0.41) | neg (2.02) | pos (0.05) | nt |
| Infant 2/M | neg (<0.05) | pos (392.6) | neg (0.27) | neg (1.33) | neg (1.44) | neg |  | neg (<0.05) | pos (95.7) | neg (0.29) | neg (1.79 | pos (0.04) | nt |
| Infant 3/F | neg (<0.05) | pos (448.5) | neg (0.24) | neg (1.37) | pos (0.39) | neg |  | neg (<0.05) | pos (413.4) | neg (0.32) | neg (1.73) | pos (0.10) | nt |
| Infant 4/F | neg (<0.05) | pos (358.8) | neg (0.11) | neg (0.04) | pos (0.007 | 1.9 x 10^2^ |  | nt | nt | nt | nt | nt | nt |
| Infant 5/M | neg (<0.05) | pos (187.2) | neg (0.29) | neg (1.15) | neg (6.57) | 3.1 x 10^2^ |  | neg (<0.05) | pos (>1000) | neg (0.30) | neg (1.77) | pos (0.05) | nt |
| Infant 6/M | neg (<0.05) | pos (>1000) | neg (0.11) | neg (1.73) | neg (1.25) | neg |  | neg (<0.05) | pos (274.1) | neg (0.28) | neg (1.78) | pos (0.06) | 7.9 x 10^2^ |
| Infant 7/M | neg (<0.05) | pos (331.5) | neg (0.11) | neg (1.72) | pos (0.67) | 2,1 x 10^3^ |  | neg (<0.05) | pos (83.1) | neg (0.27) | neg (1.74) | pos (0.06) | neg |
| Infant 8/F | neg (<0.05) | pos (33.8) | neg (0.29) | neg (1.82) | pos (0.75) | 9.1 x 10^2^ |  | neg (<0.05) | pos (176.0) | neg (0.38) | neg (1.80) | pos (0.07) | nt |
| Infant 9/F | neg (<0.05) | pos (313.6) | neg (0.28) | neg (1.63) | neg (6.42) | 2.9 x 10^2^ |  | neg (<0.05) | pos (179.9) | neg (0.43) | neg (1.96) | pos (0.07) | nt |
| Infant 10/F | nt | nt | nt | nt | nt | nt |  | nt | nt | nt | nt | nt | nt |
| Infant 11/F | neg (<0.05) | pos (45.7) | neg (0.24) | neg (2.01) | pos (0.07) | 9.9 x 10^2^ |  | neg (<0.05) | pos (242.1) | neg (0.27) | neg (2.06) | pos (0.03) | nt |
| Infant 12/M | neg (<0.05) | pos (84.2) | neg (0.34) | neg (1.81) | pos (0.05) | nt |  | neg (<0.05) | pos (54.2) | neg (0.34) | neg (1.96) | pos (0.05) | nt |
| Infant 13/F | neg (<0.05) | pos (608.7) | neg (0.08) | neg (1.44) | neg (1.37) | nt |  | neg (<0.05) | pos (119.7) | neg (0.12) | neg (1.67) | neg (2.02) | 9.2 x 10^2^ |
| Positive/  total tested | 0/12 | 12/12 | 0/12 | 0/12 | 7/12 | 6/10 |  | 0/11 | 11/11 | 0/11 | 0/11 | 10/11 | **2/3** |

*Abbreviations:* M, male; F, female; neg, negative; pos, positive; COI, cut-off index; vge, virus genome equivalent or copy number; nt, not tested.

^a^ Evaluated by Cobas E601 electrochemiluminescence immunoassay system (Roche Diagnostics, Basel, Switzerland). HBsAg, negative when < 0.05 IU/mL; anti-HBs, negative when < 10 mIU/mL; HBeAg, negative when < 1.0 COI ; anti-HBe, negative when ≥ 1.0 COI; and anti-HBc, negative when ≥ 1.0 COI.

^b^ WHV DNA evaluated by research nPCR/NAH with sensitivity 5-10 vge/mL
